# A probabilistic approach for the study of epidemiological dynamics of infectious diseases: basic model and properties

**DOI:** 10.1101/2022.08.16.22278844

**Authors:** José Giral-Barajas, Carlos Ignacio Herrera-Nolasco, Marco Arieli Herrera-Valdez, Sergio I. López

**Author notes:** Authors with equal contributions (José Giral-Barajas, Carlos Ignacio Herrera-Nolasco).

## Abstract

The dynamics of epidemiological phenomena associated to infectious diseases have long been modelled with different approaches. However, recent pandemic events exposed many areas of opportunity to improve over the existing models. We develop a model based on the idea that transitions between epidemiological stages are alike sampling processes. Such processes may involve more than one subset of the population or they may be mostly dependent on time intervals defined by infectious or clinical criteria. We apply the model to simulate epidemics and obtain realistic case fatality ratios. We also analyse the impact of the proportion of asymptomatic of infected people in the distribution of the total infected population and define a basic reproductive number, which determines the existence of a probabilistic phase transition for the pandemics dynamics. The resulting modelling scheme is robust, easy to implement, and can readily lend itself for extensions aimed at answering questions that emerge from close examination of data trends, such as those emerging from the COVID-19 pandemic, and other infectious diseases.

## 1 Introduction

The urgency to understand the population dynamics during the COVID-19 pandemic propelled new questions in many scientific fields. Analysis of clinical and experimental data has also unravelled some nontrivial aspects of the pandemic.

In parallel, many modelling efforts have attempted to provide quantitative predictions of the evolution of the pandemic, and some yielded reasonable qualitative explanations. However, the COVID-19 pandemic also unveiled some of the shortcomings of the existing modelling tools, and shed light on the fact that the construction hypotheses for many existing models are too limiting to study basic aspects of an epidemic.

We propose a new, simple model for the dynamics of infectious diseases with the idea of being able to answer basic questions and also, provide a basic paradigm from which we can draw more detailed explanations for epidemiological phenomena. The model combines ideas from sampling processes with classical approaches from dynamical systems. Of note, restricting some of the construction hypotheses of our model gives the long-established, classical SIR model (Kermack and McKendrick, 1927).

### 1.1 Epidemiological dynamics and modelling

The epidemiological dynamics of infectious diseases result from a combination of non-linear and random phenomena occurring in different levels of biological organisation. A variety of theoretical approaches, of which the most notable is the family of SIR-type models (Brauer et al., 2019; Kermack and McKendrick, 1927) has proven useful to study different aspects of epidemics for a range of diseases. Deterministic models from the SIR family are, by construction, unable to capture some aspects of an epidemic in small populations. SIR-like models, in its basic form, fail to reproduce case-fatality ratios agreeing with data. However, SIR family models can be modified to include multiple stages of infection. In these modified SIR models deaths occur only at the last epidemiological state, that is, quick deaths can only happen after an infection reaches its worse stages. Examples of these models, called *multi-scale models*, can be found in the work of Hart et al. (2020), where influenza A is taken as a case study and the error of the model’s predictions is measured in terms of the size of the data, using ad hoc synthetic patient-level data. Hart et al. worked on another multi-scale model, where early, middle, and late infection stages are included in the modelling framework to obtain accurate forecasts for ebola infections, where symptoms become more severe as infection progresses. Another example of the versatility of the SIR-like models is given in (Gaeta, 2020) where asymptomatic and non-asymptomatic compartments are included in order to evaluate the impact of the an asymptomatic population in the computation of the basic reproductive number. A novel adaptation of the SIR model can be found in (Doenges et al., 2023) where households structure for the spreading of the disease is taken into account to derive a system of ODEs to compute the basic reproduction number, the prevalence, and the peak of an infection wave.

There also is class of stochastic models, which are capable of describing other aspects of the dynamics in epidemics. The early work of Greenwood (1931) analyses the pertinence of different probabilistic models to explain mechanisms of disease spreading within households. The number of cases of the disease registered in a period of time at each household is used as input for the models, hence having the possibility of measuring the fit of each model to real data. More recently, Greenwood and Gordillo (2009) review compartmental stochastic models, focused on the distribution of the final size of the epidemics, the stochastic periodicity of the number of infectious individuals, and the (random) time that an individual remains infectious after having been infected. The model by Tuckwell and Williams (2007) uses Markovian infection dynamics that occur as binomial processes within a constant population over discrete time. Some surveys about similar stochastic models can be found in the work of Allen (2017) and Britton (2010).

Yet another approach to study epidemiological dynamics is given by the so-called *agent-based* modelling. The main idea is to construct *agents* which emulate individuals and follow a given set of rules (which can evolve differently in time, space and type of individual) of interactions between them. Then a computational methods are used to simulate the interaction of these agents and be able to observe the global dynamics and answer questions by varying the rules or the parameters used in defining them. This approach, used widely in complexity theory, allows us to include a variety of characteristics related to the epidemic which are difficult to be included in theoretical mathematical models.

Between many other examples, this approach can be used to emulate structures of social networks (as a household) with specific interaction between people (Bissett et al., 2021). In that work, a georeference system is used where the interaction between agents in the city of Burnaby, Canada (Perez and Dragicevic, 2009). To study the etiology of the *obesity epidemic* by defining agents which attempt to reproduce the complex behaviour of vulnerable people to this disease, as suggested in (El-Sayed et al., 2012). A review of the mathematical aspects of agent-based modelling can be found in (Bissett et al., 2021). Another review focused in the state-of-art in (Silverman et al., 2021), and a recent review of COVID-19 specific models is presented in (Alsharhan, 2021).

Many of the existing deterministic and stochastic classes of epidemiological models based on equations, do not offer transparent links between the mathematical expressions in the models and the mechanisms they are supposed to capture macroscopically.

Significant improvements can be made in the study of epidemiological dynamics by replacing one or more of the phenomenological construction postulates of the most basic SIR model (e.g. homogeneous mixing). This can be done by making assumptions and deriving expressions explicitly based on physical, biological, or biochemical principles underlying the infection or disease, however macroscopic.

Another issue that is often ignored is the difference between an infectious process, typically detected by laboratory tests, and the evolution of the disease possibly caused by that infection, which may only be detected if symptoms develop. This is also a problem from an epidemiological standpoint because these two phenomena are usually lumped together in construction hypotheses for models (see Allen (2008)), thus limiting the scope and the resulting dynamics (Shayak and Sharma, 2021).

Also, the clinical manifestations possibly caused by an infection are not necessarily detected or sought for. A relevant time window in this regard is the incubation period, which is asymptomatic but possibly infectious. The number of asymptomatic cases may be the main underlying cause of large epidemic outbreaks (Gerba, 2009). As noted by (Montoya et al., 2013) this dependence can be difficult to model, for example, in dengue epidemics the proportion of asymptomatics can vary across the years. Examples of communicable diseases of current importance due to their enormous burden on economic, social, and health systems, that can be asymptomatically transmitted, and therefore difficult to keep under control include malaria (Bousema et al., 2014), dengue (Grange et al., 2014), influenza (Cohen et al., 2021), COVID-19 (Gandhi et al., 2020), AIDS (Hollingsworth et al., 2008), among many well-documented examples.

The discussion above underscores the necessity of constructing models that explicitly make distinctions between infectious and illness states, or to explicitly distinguish between the exposure of susceptibles and the exposure of infected. On the other hand, agent-based modelling offers great versatility but, since mathematical analysis is not possible, it is not easy to derive general and robust explanations about the studied phenomenon. As noted by Hunter et al., it is frequently difficult to compare results between agent-based models and it is even necessary to create a taxonomy for them.

As a starting step, in this paper we present a modelling framework based on a simple dynamical principle: individuals in a given epidemiological stage may switch to a different stage through sampling. For instance, at each point in time, people who become infected are sampled from the susceptible population, and they may progress into illness, if they get sampled again.

We start by giving basic macroscopic descriptions for the transmission of infectious diseases, and describe the switching between epidemiological stages in terms of different underlying sampling processes. We then proceed to present a simple model for epidemiological dynamics, hereby called sED. Once the construction of the basic model is explained, we demonstrate a few of its mathematical properties and illustrate some of its advantages over classical epidemiological models, including the possibility of modelling infection spread in small populations with integer state variables. We also show that the sED model recovers different types of deterministic SIR models under particular sets of assumptions, and illustrate the application of the sED model with specific examples. We then show how to extend the proposed modelling framework. This extension allow us to present applications such as a detailed study of the case fatality ratio in different scenarios, an analysis of the influence of asymptotic cases in the pandemics evolution, and a extension of the basic reproductive number which value determines if we have an uni-modal or a bi-modal probability distribution for the case fatality ratio. We finish the article by presenting some discussion of the results and some future perspectives.

## 2 Modelling framework for stochastic epidemiological dynamics

### 2.1 Working definitions and conceptual distinctions

*Susceptible* individuals are those that can be infected. *Exposed* individuals are those in contact with the pathogen. Exposure may change over time depending on different factors like physical distancing policy, personal mobility, the time of the day, etcetera. Naturally, only susceptible individuals that are *exposed* can become infected, and an individual is *infected* if the pathogen finds a susceptible tissue and starts replicating. After some time in which the pathogen replicates, infected individuals become able to transmit the pathogen and start contributing to the chain of transmission. The time relative to the (initial) inoculation is a random variable (Fenner et al., 1987). The *incubation* time can be regarded as the time interval between the start of replication and the start of pathological processes associated with pathogen invasion (Li, 2010, p.25). However, since symptoms are usually the way in which pathology is detected, the incubation time is usually referred to in medical epidemiology as the time lapse between the suspected time of infectious contact and the start of symptoms (Dicker C. et al., 2006; He et al., 2020). This is typically a problem since infected people may not be symptomatic (they do not show obvious symptoms), as their infection progresses. In the case that disease ensues, the severity may progress from mild to worse until recovery or death.

The time individuals spend being infected is a random variable that often is modelled as a Gamma-distributed random variable (Anderson and Watson, 1980; Feng et al., 2007; Lloyd, 2001; Vazquez, 2021). For practical purposes, the infection time can be thought of as a sum of an initial replication period of the pathogen causing the infection, and a second period during which an individual may have detectable symptoms. The individual may be infectious shortly after the first pathogen invasion, during the incubation period, and before symptoms appear (e.g. COVID-19). This can be modelled by dividing the infectious period into several stages. After the infection period, individuals may develop other phases of the disease that are not necessarily infectious. For instance, in both symptomatic and asymptomatic COVID-19, the initial viral phase might be followed by an inflammatory phase (García, 2020; Manjili et al., 2020) where the probability of infectious virus shedding decreases dramatically (van Kampen et al., 2021). A longitudinal study of people infected with SARS-CoV-2 (Boucau et al., 2022) suggests the possibility of shedding virus for more than 5 days (possibly up to 15 days) after symptom onset. In the best-case scenario, the immune system develops a protection profile against the pathogen and the individual recovers soon after that. In a worse-case scenario, the response of the immune system may be too strong, inflicting damage on different tissues, exacerbating the pathology further, causing severe illness and potential death. Alternatively, it is also possible that individuals are born immune or become immune. This may happen for reasons that include having beneficial mutations in their genome (Allers and Schneider, 2015), having acquired cross-immunity upon exposure to other pathogens (Doolan et al., 2009), or after vaccination (Clem, 2011).

In consideration of the observations made above, and others mentioned further in the perspectives subsection of the discussion, we developed a hierarchy of models for stochastic epidemiological dynamics referred to as sED models from here on, that can be extended depending on the level of detail sought and the questions of interest.

### 2.2 Modelling rationale

We regard any epidemiological classification of individuals from a population as a dynamical partition into subsets that change in time. The subsets that make up the partition can be represented as nodes in a directed graph with nodes and edges can be respectively weighed according to the size of the population they represent, or alternatively, the probability that an individual changes to the epidemiological stage at the end of the edge (Fig. 1).

**Figure 1:**
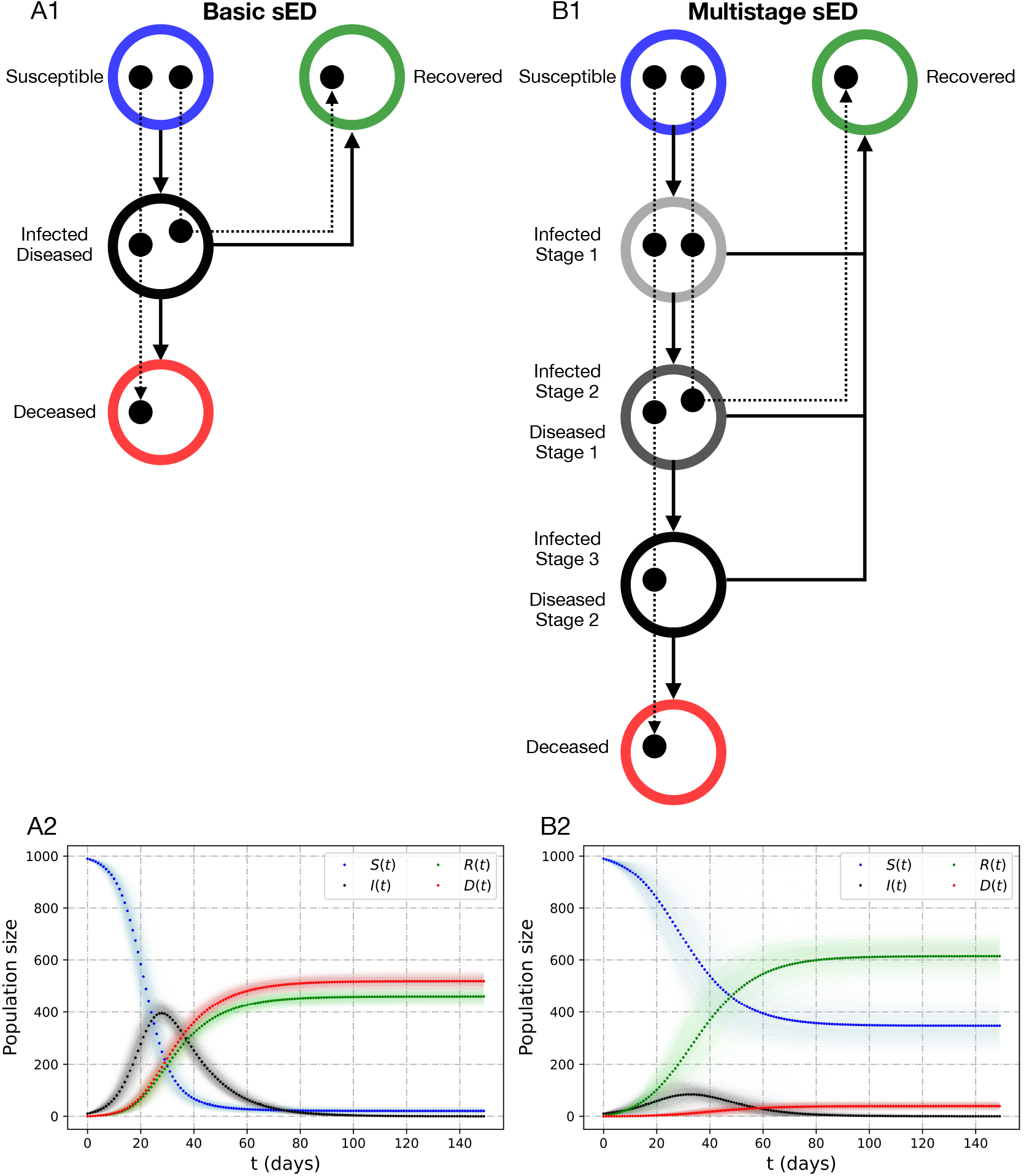
Different dynamical scenarios contemplated by the sED modelling scheme outlined in A1 and B1. The light shades in A2 and B2 represent the confidence bands generated with 2,000 simulations and the solid dots are taken as the average of those simulations. The dynamics in the two models are defined as having the same average time for recovery and death. However, they display different time courses for infection and final outcome. See tables 1 and 2 for further information.

The dynamics for this model are based on two simple but powerful concepts.

- First, each infection can be thought of as a *one-way, path* starting at the node representing the susceptible subpopulation, and ending at one of the terminal nodes in the graph representing the recovered state or death due to infection.
- Second, the epidemiological state *s*(*t*) of any individual may change during the time interval [*t, t* + *h*] with a certain probability 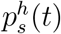, and remain the same with probability 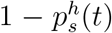. Such changes are assumed to occur independently between individuals.

#### 2.2.1 Epidemiological sampling and state switching

The epidemiological state *s* of an individual at time *t* + *h* can be thought of as the result of a *routing* process in which individuals are subjects in *Bernoulli* trials occurring between times *t* and *t* + *h*, with probability of success (switch) 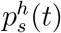. The number of people that change their epidemiological state between times *t* and *t* + *h* is thus the number of successful trials in the *Bernoulli* sample drawn at time *t*. It is reasonable to assume independence between switching events for different individuals. Within a homogeneous population (e.g. similar co-morbidities), every individual can be assumed to have the same probability of switching between epidemiological states, in which case the number of people who switch can be regarded as a *Binomial random variable* in which the number of trials is the number of individuals initially considered. If there is more than one possible new epidemiological state to switch to, 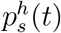 can regarded as a sum of the probabilities of switching to the new different states, and the individuals that switch to the different states can be thought of as a *Multinomial* sample with a number of trials equal to the number of people in state *s* at time *t*. The epidemiological dynamics of a population formed by different, but relatively homogeneous subpopulations can then be modelled by keeping track the individual changes in epidemiological state, and updating the number of people in the different stages after each time step.

Reasonable approximations for *populations* formed by heterogeneous groups can then be derived by assuming homogeneity among individuals within a group and tracking the evolution of the different groups. For instance, in some cases it would be reasonable to assume that the (population) average probabilities for switching describe the population dynamics at large, and therefore, sampling for those individuals switching epidemiological states can be done in each subpopulation by performing independent Bernoulli trials, assuming that success occurs with a probability equal to the average probability of switching between epidemiological states for that subpopulation.

The first model we present will be based on this basic assumption which implies two sampling processes on one homogeneous population: one from the susceptible population that depends on contacts between susceptible and infected, and another from the infected that only depends on time from the start of infection (Fig. 1 A, left column).

#### 2.2.2 Time-dependent switch in epidemiological state

Let *τ*_*s*_ be a random variable representing the waiting time for a switch to a new epidemiological state *s*. For instance, *τ*_*R*_ represents the expected time for recovery for an individual at some infectious state. The probability of switching within *h* time units can then be written as 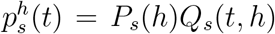 where *P*_*s*_(*h*) represents the probability that the switch occurs within *h* time units, and *Q*_*s*_(*t, h*) represents the probability that the conditions that are necessary for the switch occur at time *t*. Assuming that the time *τ*_*s*_ for a switch from the epidemiological state *s* is an exponential random variable with mean waiting time *μ*_*s*_,

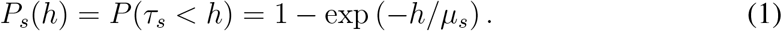

We now proceed to show the derivation for the basic version of the stochastic model for epidemiological dynamics, which is capable of capturing different aspects of epidemiological dynamics, showing some of their properties and advantages over classical models. We do so taking into account the characteristics and data reported for COVID-19.

#### 2.2.3 The start of the chain of transmission: switching from susceptible to infected

Suppose that every member of a population is either susceptible to infection, non-susceptible, or infected by a certain pathogen (e.g. SARS-CoV-2), at any point in time. Let non-negative integers *T*(*t*), *S*(*t*), and *I*(*t*) represent the sizes of the whole population, those susceptible to infection, and those infected at time *t* (in days). *T*(*t*) − *S*(*t*) is the size of the non-susceptible population at time *t, i*.*e*. those that do not participate in the chain of infections.

Assume that infections are *independent* among individuals, with an average probability of infection *p*^*h*^(*t, I*) that depends on factors that include the time of exposure *h* and the available inoculum, which can be thought of as a monotonically increasing function of *I*. In addition, suppose that each of the susceptibles can be infected within *h* time units with probability *p*^*h*^(*t, I*), or remain susceptible with probability 1 − *p*^*h*^(*t, I*). The number 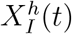 of newly infected people (absolute incidence) can then be thought of as a *Binomial* random variable sampled from *S*(*t*) individuals. One possibility is to define

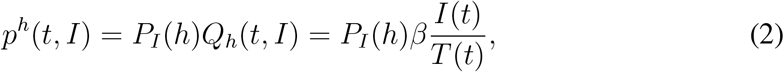

where *β* is the probability that a susceptible person is infected after having an infectious contact with the infectious pathogen, *I*(*t*)*/T*(*t*) represents the proportion of infected individuals within the population at time *t*, and *P*_*I*_(*h*) represents the probability that the infection occurs within a time window of length *h*, while being *exposed* to the infectious pathogen. It its worth mentioning at this point that the inoculum may also depend on the age of infection of individuals, which may not be easily characterised. The inoculum may include sources other than the infected individuals. To take this into consideration, one could give more precise definitions of *Q*_*h*_(*t, I*), however this is out of the scope of this work. The number of newly infected individuals between times *t* and *t* + *h* can then be thought of as a random variable 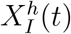 with *Binomial* (*S*(*t*), *p*^*h*^(*t, I*)) distribution, and the expected number of newly infected individuals would then be *p*^*h*^(*t, I*)*S*(*t*). The number of susceptible people at a time *t* + *h* can then be calculated as,

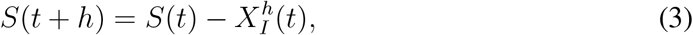

which means that the expected change between *S*(*t*) and *S*(*t* + *h*) is a decreasing function of *t*, for all *h >* 0. As a working definition, we will consider the start of an epidemic when the expected number of infections, *S*(*t*) · *p*^*h*^(*t, I*), becomes increasing as a function of time.

We now describe how to take into account the possibility of death due to disease.

### 2.3 Infections and possibly fatal outcomes

Assume that an initial number of infections *I*(0) = *I*_0_ are caused by a primal source of pathogens, and that those infections are transmissible to the rest of the population. Assume also that infected individuals shed the same (average) inoculum per unit time. For now, all susceptible and infected individuals are assumed to participate in the chain of infection without restrictions. In a later section, we will explore the effects of extending the model to explicitly include exposure factors for susceptible and infected, separately.

#### Immunity and death

For this first model, assume that some of the infected individuals die after becoming infected, and the rest recover and become immune from then on, as assumed in the classical SIR formulation. Notice that the assumptions above do not take into consideration any clinical aspects associated to the infection, with only two possible stage switches to become deceased or eventually recover. Let 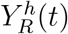 and 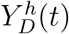 respectively represent the numbers of people that that recover or die between *t* and *t* + *h*. Those that remain infected after that time interval are then

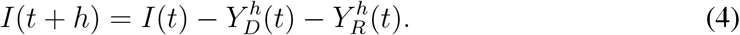

The probabilities that a person is removed from the infected group (i.e. recovers or dies), or that the person remains infected within a small time interval of length *h*, are assumed to be independent of the state of the epidemics at time *t*; thought of as to depend only on the physiological state of the individual facing the infection. Independence between infections enables the possibility of regarding 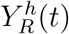 and 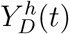 as outcomes from a *Multinomial* sample from *I*(*t*), with respective probabilities 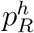 and 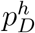, for *t* ∈ [*t, t* + *h*].

In other words, the triplet

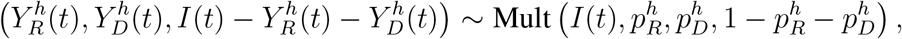

and the dynamics of the infected, recovered, and dead at time *t* + *h* can be written as a stochastic dynamical system of the form

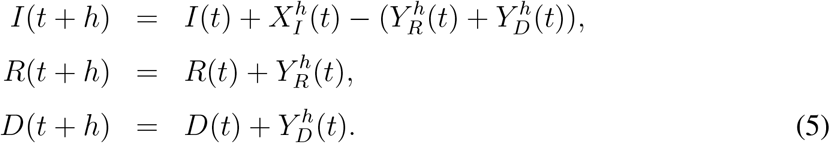

where *R*(*t*) and *D*(*t*) are non-negative integers respectively representing the number of recovered or deceased individuals at time *t*.

#### 2.3.1 Probabilities for recovery and death

Let *τ*_*R*_ and *τ*_*D*_ be positive-valued random variables respectively representing the waiting time for an infected person before recovery or death. Bearing in mind that the distributions of *τ*_*R*_ and *τ*_*D*_ may vary substantially depending on the pathogen (Baron, 1964), assume that infected individuals can either clear a pathogen within an average time *μ*_*R*_, or alternatively, the individual may die from disease caused by the infection after an average time *μ*_*D*_. The probabilities that a person is removed from the infected group or that remains infected within a small time interval of length *h* can be estimated by assuming *Geometrically* distributed times for recovery and death, so that

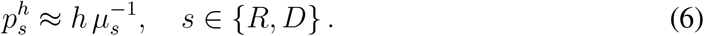

with 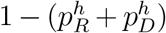 representing the probability of remaining infected within *h* time units. Large enough values of *I*(*t*) yield a *Poisson* approximation of the form

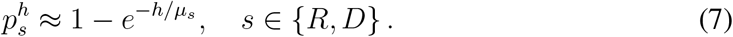

Take for instance the epidemiological dynamics of COVID-19 (Fig. 2). Data from the first symptomatic cases of COVID-19 during 2020 in China suggested that deaths in hospitalised individuals occurred between 2 and 3 weeks after start of symptoms (Zhou et al., 2020). In contrast, the hospitalisation time for survivors was reported to be between 3 and 4 weeks (Zhou et al., 2020). Also, the average incubation time was reported to be between 4 and 6 days (Quesada et al., 2020), and during this interval of time time those infected could shed inoculum while remaining asymptomatic. Taking the above observations into account, we assume values for *μ*_*R*_ and *μ*_*D*_ to be 21 and 28 days, and 28 and 35 days, respectively to construct the basic sED model with dynamics illustrated in figure 2.

**Figure 2:**
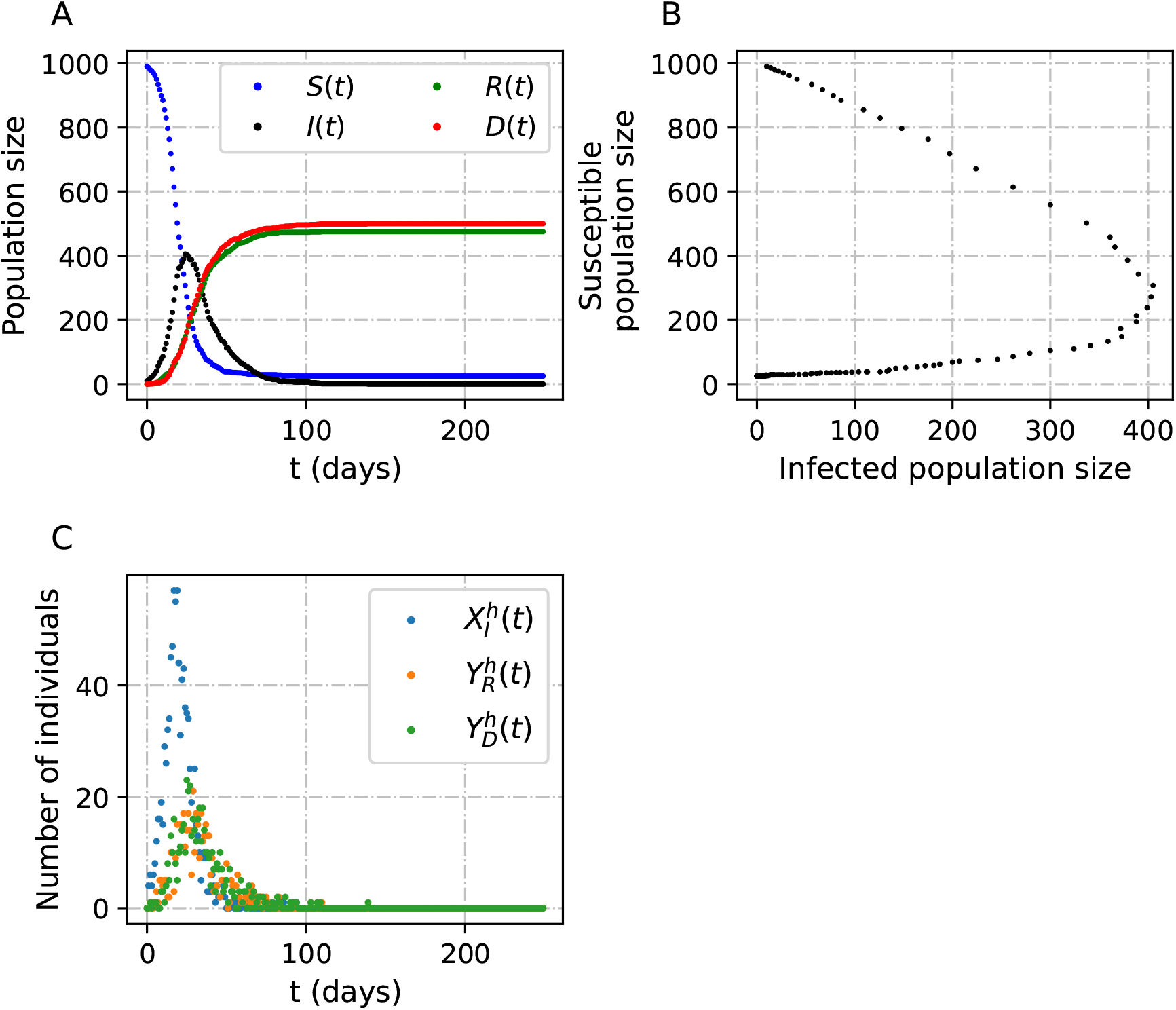
Dynamics and geometry of the sED model. (A) Epidemiological curves obtained with the sED model. (B) *I* − *S* phase plane. (C) Numbers of newly infected individuals, of people that recover, and of people that die over time. The parameters used in this simulations were *μ*_*R*_ = 27, and *μ*_*D*_ = 24. Other parameters can be found in table 1.

The joint dynamics of the decreasing size of the susceptible population and the probability of infection result in a sequence of probability mass functions for the newly infected that have many remarkable features (Fig. 3). First, the peak probability has a “U” shape as a function of time, taking the highest values at the beginning and end of the epidemic, and the lowest at the peak of infections. The number of newly infected that corresponds to that peak probability has the opposite behaviour, starting and ending at very low values, and reaching a maximum at the peak of the epidemic. This is in line with the idea that unless the value of *β* is very large, the probability that a large number of new infections occur is low, at any point in time, regardless of the population size.

**Figure 3:**
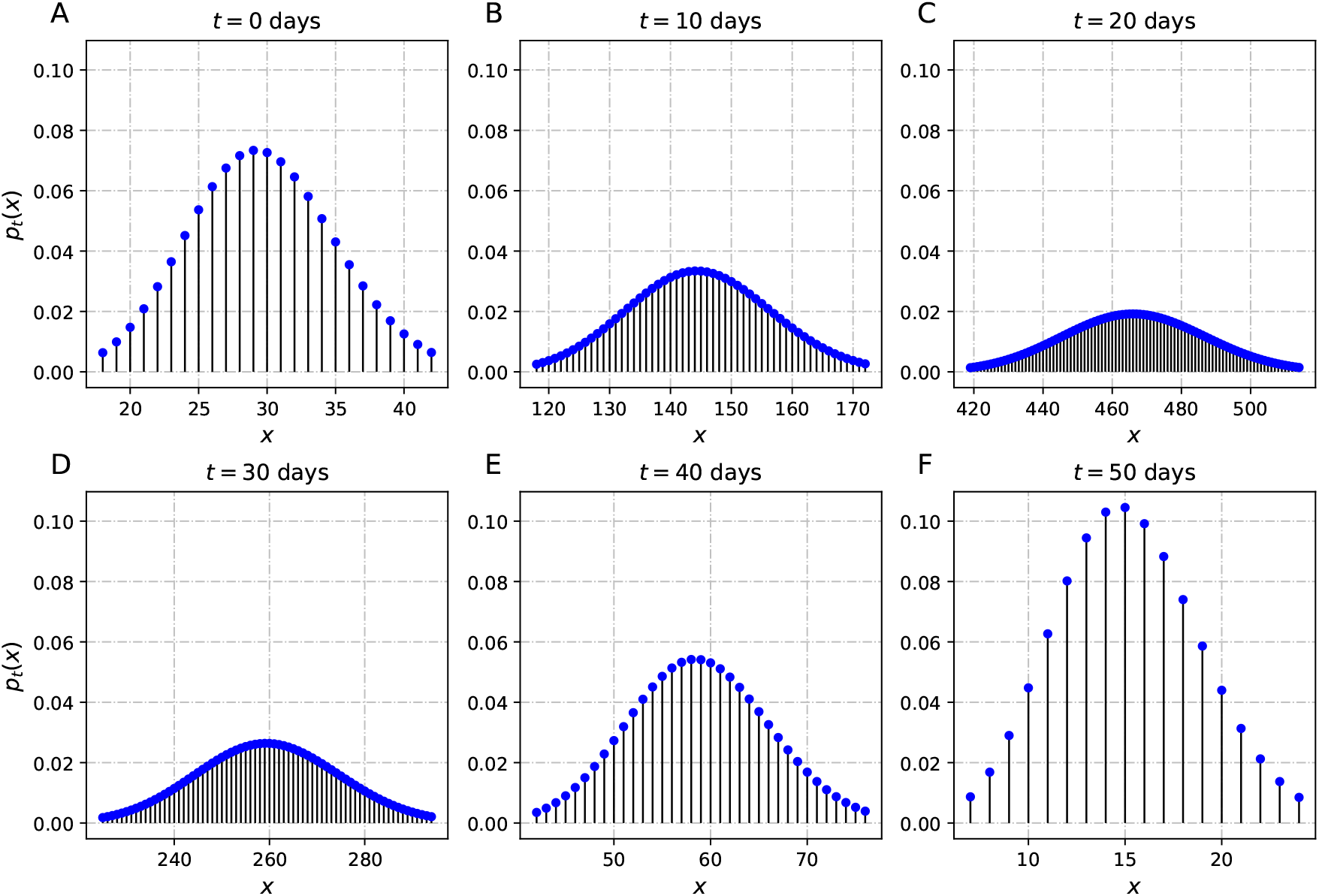
Example of the evolution of the probability mass function *p*_*t*_(*x*) of new infections *x* in the basic sED model as a function of time *t*. Time progresses through the panels, where panel A represents the beginning of the epidemic, panel B represents day 10, panel C represents day 20, and so on until panel F representing day 50. The parameters used in the simulation were *T*_0_ = 10, 000, *β* = 0.3, *μ*_*R*_ = 27, *μ*_*D*_ = 24 and *h* = 1.

Before presenting applications and further extensions of this basic model, we demonstrate some mathematical properties derived directly from equations (3)-(6).

#### 2.3.2 The routing probability and the mean infection time

We first answer a fundamental question: What is the probability that a given infected person recovers?

The multinomial trial with probabilities 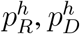, and 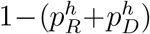 for any time *t* is independently repeated for time windows of size *h*, for as long as the population has infected individuals. The event in which an individual *stops being infected* can then be thought of as a sequence of independent trials, each lasting *h* time units, in which the individual remains infected and in the last trial the person either recovers or dies. The *routing probability* to the recovered state, that is, the probability that an individual eventually recovers, is then given by

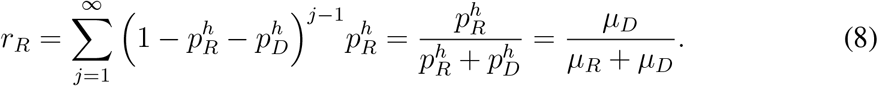

The probability that an infected individual eventually dies is *r*_*D*_ = 1 − *r*_*R*_. Explicitly,

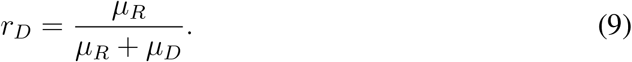

Note that these two probabilities do *not* depend on the time step *h*.

The dynamics of the multinomial trials for each infected person can also be thought of as Bernoulli trials with probability of success *p*_*R*_ + *p*_*D*_, and the elapsed time *τ*_*I*_ can then be thought of as a random variable with a mean 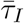 given by the inverse of the probability (rate) of leaving the infectious state,

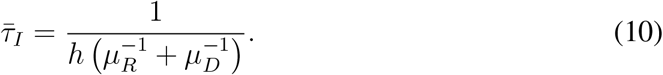

This means that the model can be formulated in terms of the routing probabilities and the mean time spent in the infection stage, 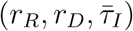, which could possibly be obtained from hospitalisation data, instead of the mean infection times spent by a person before recovery or death, (*μ*_*R*_, *μ*_*D*_). Using equations (8)-(10) we have

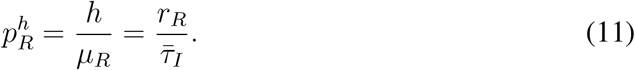

The inverse of the mean time spent in the infected phase is thus proportional to the probability that within *h* time units an individual stops being infected, regardless of the outcome.

### 2.4 Continuous time Markov process approximation

Now consider an approximation for the simple sED model, in the case when the step size *h* (the discrete units of time for the simple sED model) decreases to zero for integer population sizes. The approximation gives insight about the theoretical properties in the limit for small step sizes, and the limit process is relatively easy to simulate since it is a continuous time Markov process (Fig. 4).

**Figure 4:**
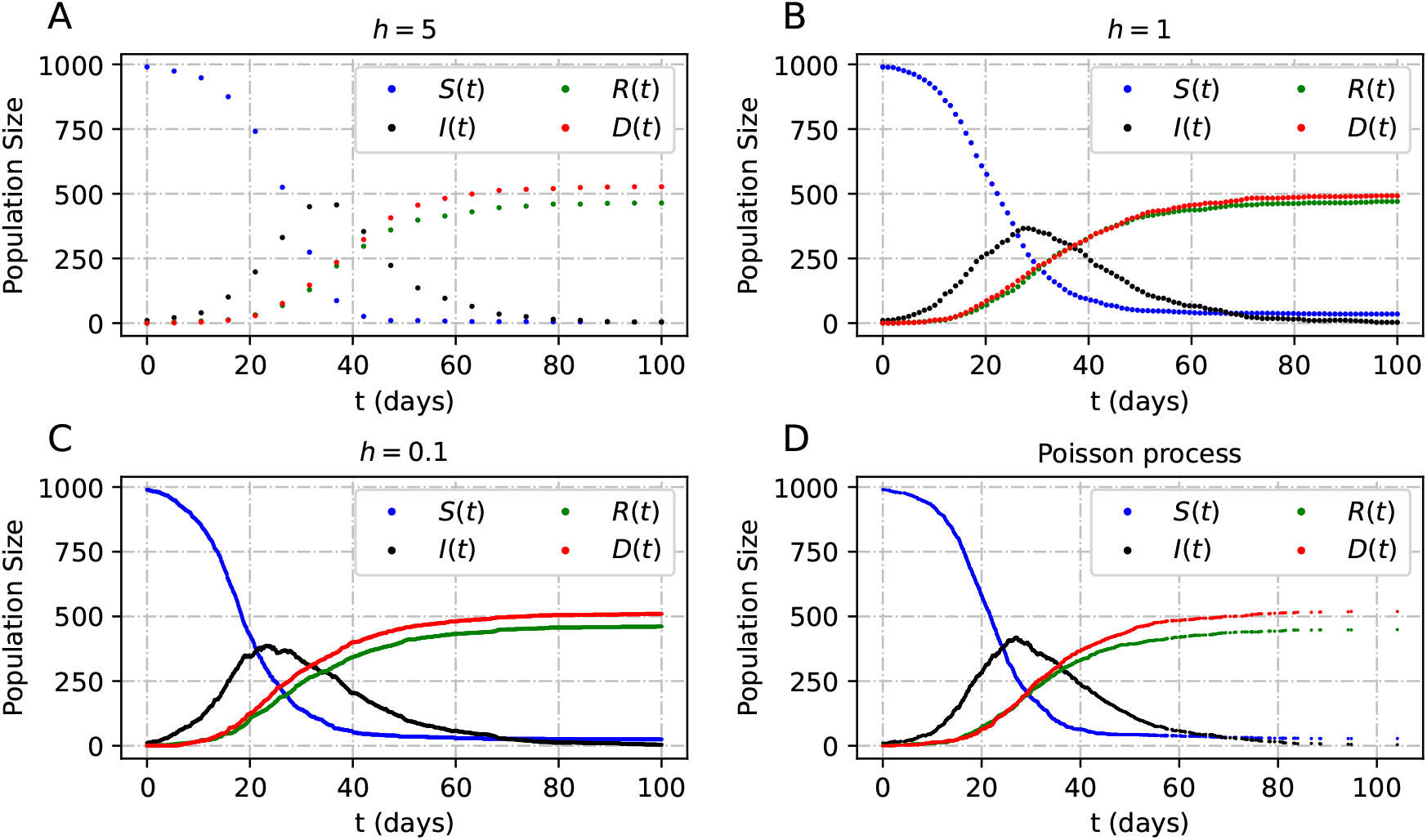
Dynamics of the sED model for decreasing step sizes (*h*) (A-C), and the approximation of the model using the Poisson process (D). For this simulation *μ*_*R*_ = 27 and *μ*_*D*_ = 24, and the other parameters can be found in table 1.

**Figure 5:**
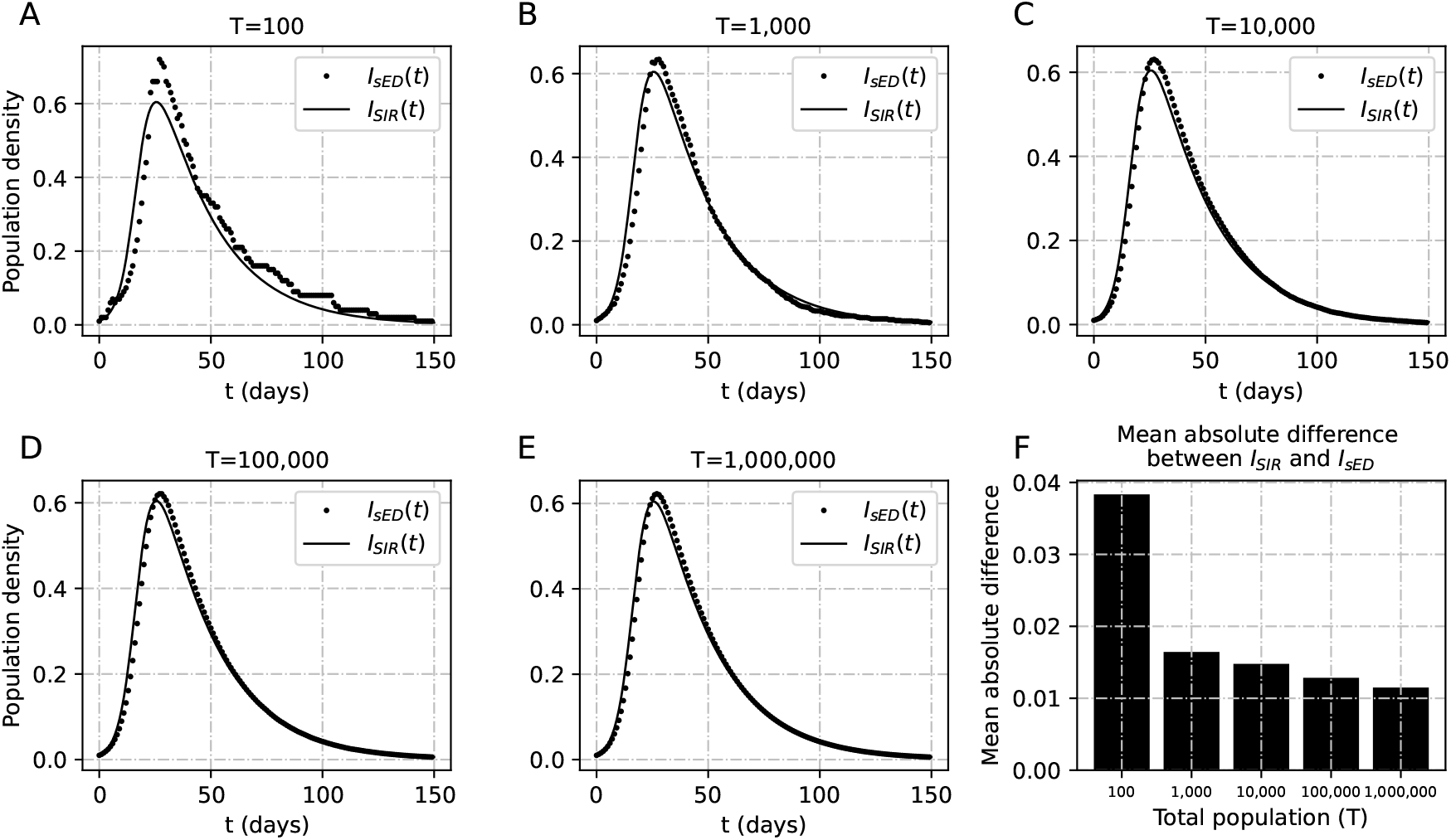
Density of infections for different population sizes of SIR vs sED dynamics obtained by replacing the sampling processes by their mean at each time step (A-E). The bars on panel F indicate the size of the difference between the total infected over time between the two models as a function of the population size. For these realisations, *μ*_*R*_ = (27 + 24)*/*2 for the sED model without deaths, and *β* = 0.3 and *γ* = 2*/*(27 + 24) for the SIR model.

**Figure 6:**
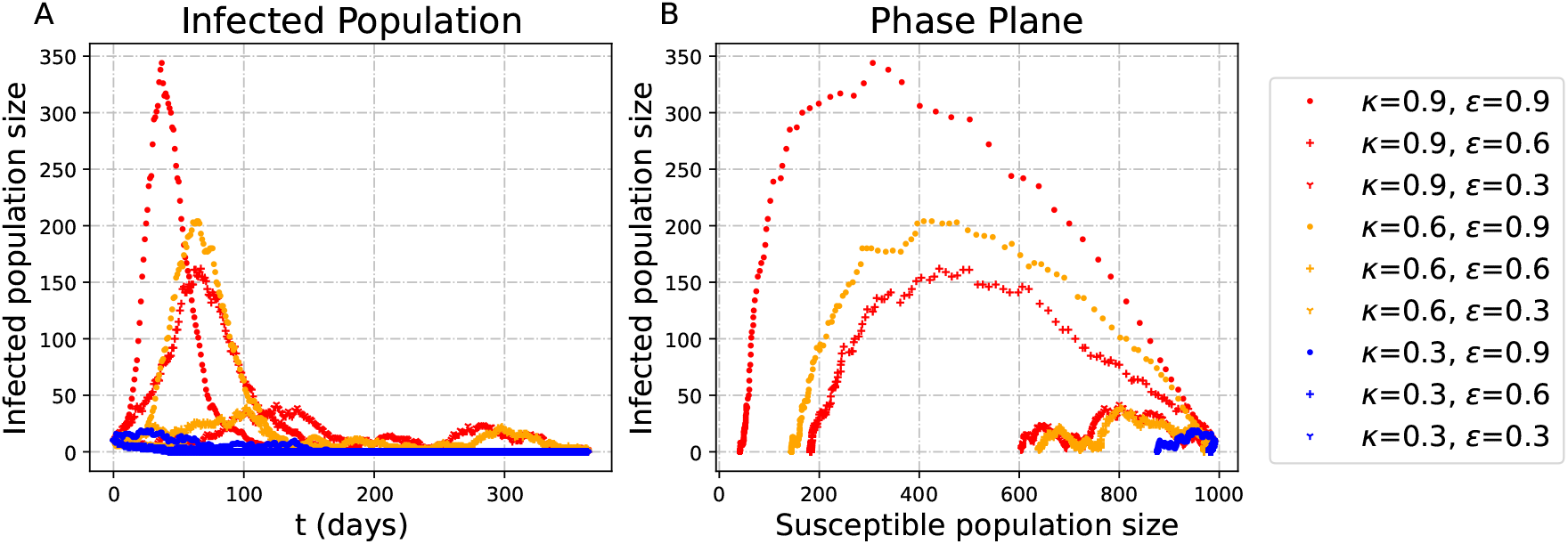
(A) Infection processes varying the exposure for susceptible and infected (*ε* and *κ*). (B) The *S* − *I* phase plane for different values of *p*. Realisations were calculated using *μ*_*R*_ = 27, and *μ*_*D*_ = 24.

Recall that a non-homogeneous Poisson process {*N*(*t*) : *t* ≥ 0} with intensity *λ*(*t*) and starting at the origin, can be assumed to have independent increments such that,

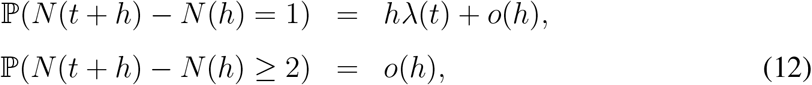

for any *t* ≥ 0 (see for instance Basu, 2003, p. 142). Consider the sED state vector {(*S*(*t*), *I*(*t*), *R*(*t*), *D*(*t*)) : *t* ≥ 0} with dynamics given by equations (3)-(5), starting from some initial condition (*S*(0), *I*(0), *R*(0), *D*(0)). Assume that

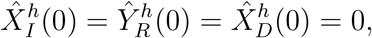

with the super index *h* emphasising dependence on the size of the time step. After *k* steps, the cumulative sampling processes can be written as

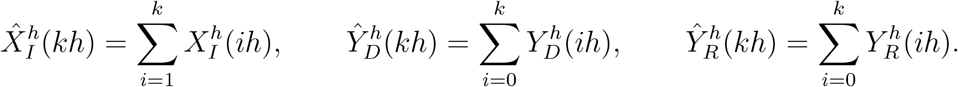

These processes are defined for a discrete set of times {0, *h*, 2*h*, …}, but they can be extended for any positive time as left-continuous step functions on continuous time, taking a discrete set of values. For example, 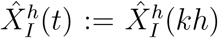 for any *kh* ≤ *t <*(*k* + 1)*h*, with *k* ∈ ℕ. As a consequence, the limit processes

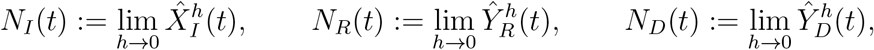

are non-homogeneous Poisson processes. We show that in the following lines.

By construction *N*_*I*_, *N*_*R*_ and *N*_*D*_ start at the origin, and the random variables 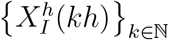 are independent, which means that the process 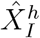 has independent increments on disjoint intervals of the form [*n*_1_*h, n*_2_*h*] where *n*_1_ *< n*_2_ are non negative integers. Therefore, the limit processes *N*_*I*_, and by a similar argument, *N*_*R*_ and *N*_*D*_, have independent increments. Further, for *t* ∈ [*kh*, (*k* + 1)*h*), *S*(*t*) = *S*(*kh*), *I*(*t*) = *I*(*kh*), and

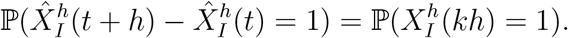

Since 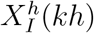 has a Binomial distribution with parameters *S*(*kh*) and *hβI*(*kh*)*/T*, respectively, it follows that

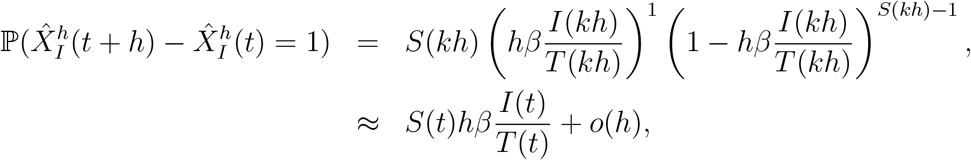

for *h >* 0. Also the probability

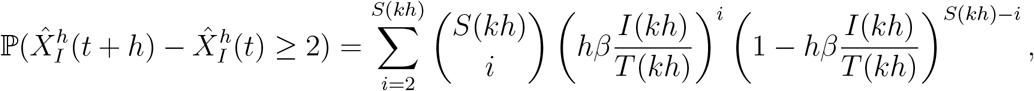

has order *o*(*h*), for *h >* 0. In the limit as *h* goes to zero, the process *N*_*I*_ satisfies equation (12) and 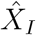 is thus a non-homogeneous Poisson process with intensity *S*(*t*)*βI*(*t*)*/T*(*t*). Since, 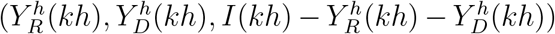 is Multinomially distributed for *kh*, we have that 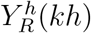 is Binomially distributed with parameters *I*(*kh*) and 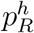. Likewise, 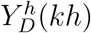 is Binomially distributed with parameters *I*(*kh*) and 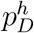. The processes *N*_*R*_ and *N*_*D*_ satisfy (12), which can be proven by following the same reasoning as for *N*_*I*_.

Taking into account the arguments just presented, as the step size *h* decreases to zero, the sED system {(*S*(*t*), *I*(*t*), *R*(*t*), *D*(*t*)) : *t* ≥ 0} based on Binomial samples converges in distribution with order *o*(*h*) to the system 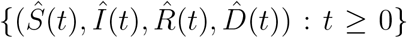 based on Poisson sampling, starting from the same initial conditions, with dynamics given by

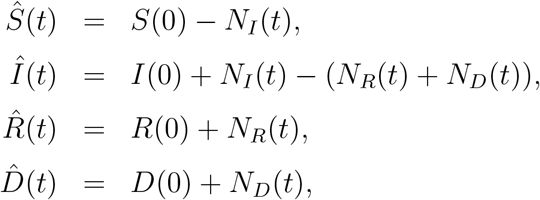

where *N*_*I*_, *N*_*R*_, *N*_*D*_ are non-homogeneous Poisson processes with corresponding intensity rates 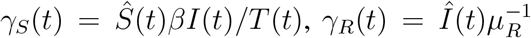, and 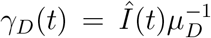. Note that the model dynamics depend on the size of the population *T*(*t*) for each time *t*. In the idealised situation where the total size of the population is constant, *T*(*t*) ≡ *T* (no deaths), the model is greatly simplified because the limit process 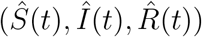 boils down to a continuous time Markov chain with state space contained in the set {0, 1, …, *T*}^3^ and infinitesimal generator matrix with transition rates given by

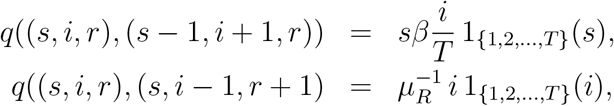

for *s, i, r* ∈ {0, 1, …, *T*}. Therefore, if total population is constant, then the Markov approximation to continuous time coincides with an stochastic SIR model previously proposed (see Section 7.6 in Allen, 2010).

The basic models presented up to this point have some advantages, including the possibility of modelling epidemics in very small populations. However, these basic models have some limitations that can be overcome by extending the basic model. For instance, we will include different stages for the severity of infection to analyse the impact they have in the case-fatality ratios, a quantity often ignored in modelling studies. Importantly, the extensions are constructed following the same rationale used for the basic models.

**Table 1:**
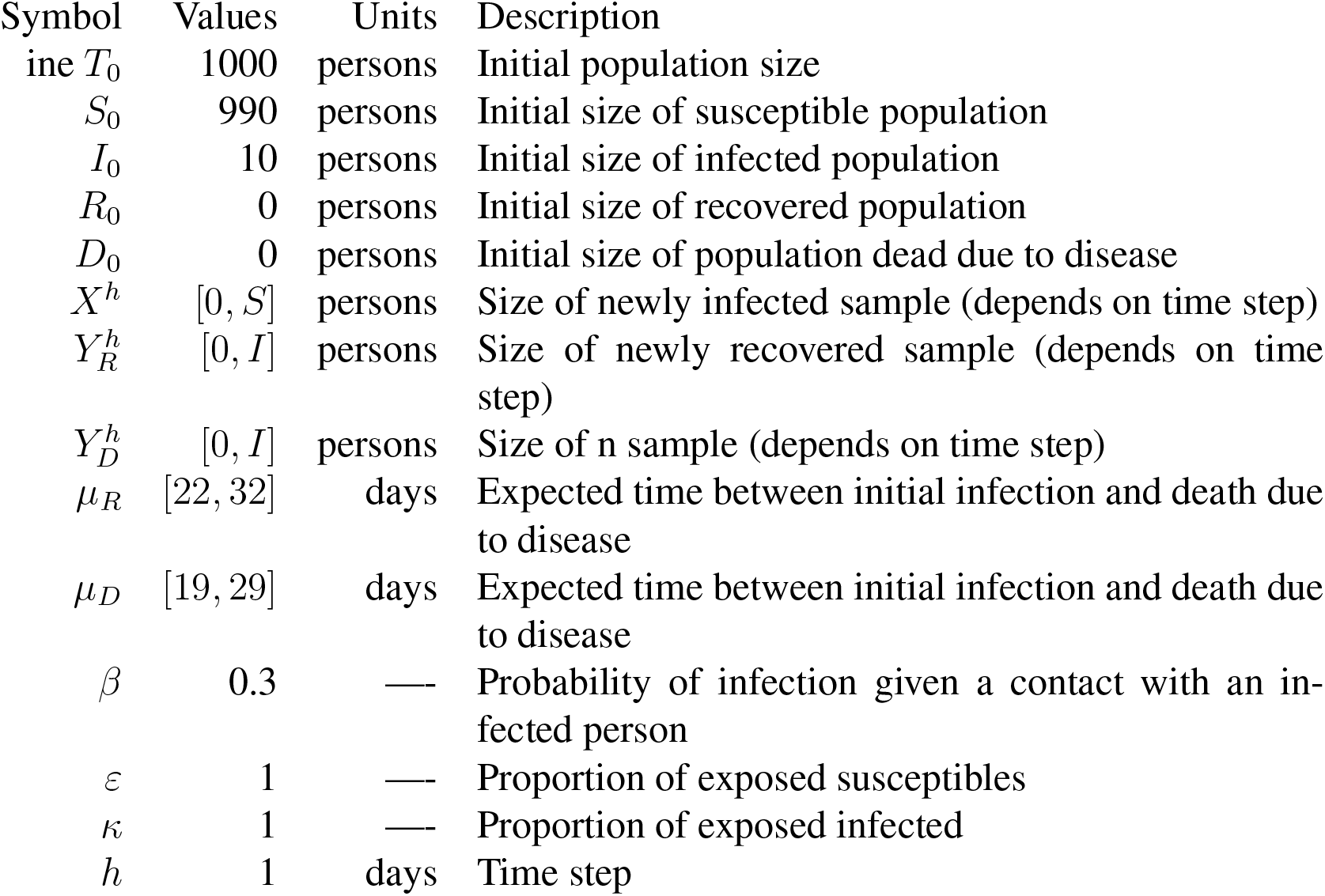
Notation and values used for simulations with the basic sED model (Li et al., 2020; Phucharoen et al., 2020; Zhou et al., 2020).

## 3 Applications

### 3.1 The classical SIR model from the basic sED model

The classical SIR model describes the dynamics of an epidemic in a large population of *constant* size, in terms of three non-overlapping subpopulations representing susceptible, infected, and those that cannot participate in the chain of infections (either formerly infected or immune from the start). One key assumption of the classical SIR model is that individuals that cease being infected do not become susceptible again. In its continuous time and continuous state version, the SIR model can be written as a system of differential equations constructed under the assumption that the population is heterogeneously mixed and of fixed size (no deceases due to infection), with transmission occurring possibly after a contacts between susceptible and infected individuals.

An analogue of the classical SIR model can be obtained as a particular case of the basic sED model (equations (3)-(5)) by assuming that: (i) there are no deaths due to infections, which makes the population size *T* a constant; (ii) the samples 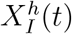 and 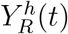 are replaced by their expected values, which yields a deterministic system of difference equations; (iii) the population size is very large; (iv) the (integer) sizes of the epidemiological classes are replaced by subpopulation densities, thus enabling the possibility of thinking about state variables as continuous; and (v) the time step can be arbitrarily small to replace the difference equations with differential equations where the densities are continuous variables changing with respect (continuous) time.

Briefly and explicitly, the expected number of new infected between times *t* and *t* + *h* is *h β S*(*t*) *I*(*t*)*/T*(*t*), which means that, on average, the absolute incidence (new cases) between *t* and *t* + *h* can be written as

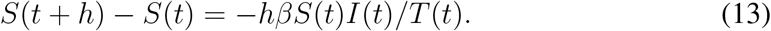

Similarly, the number of people expected to stop being infected between *t* and *t* + *h* is 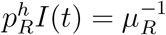, and the average change in a window of *h* time units is thus

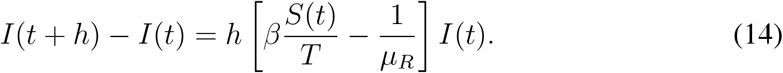

Then, if the (initial) population size *T* is large enough, we can let *x*(*t*) = *S*(*t*)*/T, y*(*t*) = *I*(*t*)*/T*, and *z*(*t*) = 1 − (*x*(*t*) + *y*(*t*)), and then write differential equations from equations (13)-(14) describing the dynamics of (*x, y, z*) by taking the limit as *h* tends to 0. If we let *∂*_*t*_ denote the instantaneous rate of change with respect to time, then the dynamics for *x* and *y* can be described by differential equations of the form

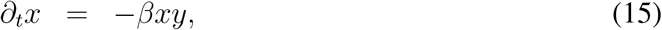

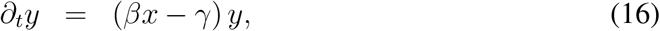

with *∂*_*t*_*z* = − (*∂*_*t*_*x* + *∂*_*t*_*y*) = *γy* representing the time-dependent change in the density of the population that is immune to the disease (*i*.*e*. does not participate in the chain of transmission), and *γ* = 1*/μ*_*R*_.

#### 3.1.1 Is it possible to write a simple and well posed extension for the continuous and autonomous basic SIR to account for deaths without further changes?

Can we change equation (16) to include the possibility of death due to disease? One way to do that would be to split *γ* as a sum of two rates, one representing recovery, the other death, respectively. As a consequence, *z*(*t*) would be now a sum of population densities *d*(*t*)+*r*(*t*) where *d*(*t*) = *D*(*t*)*/T*(*t*) represents the proportion of people dying at time *t*, and *r*(*t*) = *R*(*t*)*/T*(*t*) represents the proportion of people that do not participate in the chain of transmission at time *t* (either because they recovered or because they were immune to infection from the start). A typical setup for this would be to extend the system (15)-(16) to include

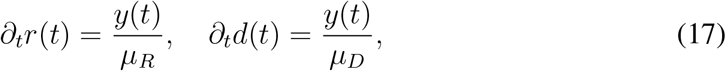

which could, in principle, be solved numerically to obtain approximations for the dynamics of the state vector (*x, y, r, d*), for each time *t*. However, the total population size at time *t* is *T*(*t*) = *S*(*t*) + *I*(*t*) + *R*(*t*), so that 1 = *x*(*t*) + *y*(*t*) + *r*(*t*) for all *t* ≥ 0, and

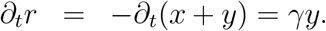

But *γy* = *∂*_*t*_*z* = *∂*_*t*_*r* + *∂*_*t*_*d*, which means that *∂*_*t*_*d* ≈ 0 for all *t* ≥ 0. As a consequence, adding dynamics for deaths due to infection would add an inconsistency in the way the model is posed.

Assuming that the total population is large and constant imposes a number of restrictions for the construction of deterministic and autonomous models for epidemiological dynamics as proposed in standard textbooks and articles alike (Allen et al., 2008; Keeling and Rohani, 2008). The above paragraphs provide some insight for one limitation that may be encountered while trying to construct possible extensions that include death due to disease. There are however, extensions that include death due to disease at the expense of also including terms we did not consider here, to guarantee consistency in the system of equations (see Section 2.2 in Keeling and Rohani, 2008).

### 3.2 Limiting the exposure of the susceptible vs. limiting the exposure of the infected

Assume that a certain proportion *ε* of the susceptible are exposed to inoculation, and also, that only a proportion *κ* of those infected actively sheds inoculum.

Suppose that at any time *t* (days), non-negative integers *T*(*t*) and *S*(*t*) represent the size of the whole population and those susceptible to infection at time *t*. Let *ε*(*t*) and *κ*(*t*) respectively represent the proportion of the susceptible population that are exposed to the pathogen at any time *t*, and the proportion of infected people that shed inoculum. The values of *ε*(*t*) and *κ*(*t*) depend on factors that include the mobility of the population and other behavioural patterns.

The number 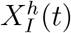 of newly infected people (absolute incidence) can then be thought of as a *Binomial* random variable sampled from ⌊*ε*(*t*)*S*(*t*)⌋ individuals. Assuming homogeneous mixing between the exposed susceptibles and the exposed infected, define

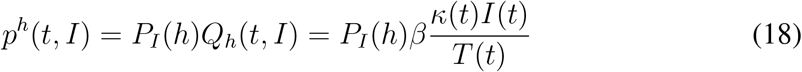

where *β, κ*(*t*) and *I*(*t*)*/T*(*t*), respectively represent the probability that a susceptible person is infected after having an infectious contact (contact with the pathogen), the proportion of infected individuals shedding inoculum, and the proportion of infected individuals within the population at time *t. P*_*I*_(*h*) represents the probability that the infection occurs within a time window of length *h*, while being *exposed* to the pathogen causing the disease of interest. As a consequence, the number of newly infected individuals between times *t* and *t* + *h* can be thought of as a random variable 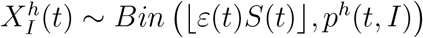, and the expected number of newly infected individuals would then be *p*^*h*^(*t, I*)⌊*ε*(*t*)*S*(*t*)⌋.

Recall that a Binomial random variable with parameters *n* and *p* converges to a *Poisson* random variable with parameter *np* as *n* tends to infinity. In this way if ⌊*ε*(*t*)*S*(*t*)⌋ ≥ 10 a Poisson random variable with parameter 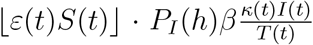, has a probability distribution very close to the one of 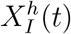 (Poor, 1991). Observe that *ε*(*t*) and *κ*(*t*) appear as a product in the parameter of the Poisson approximation. Therefore, some degree of symmetry can be expected for large enough populations in the behaviour of *I*(*t*)], and other quantities depending on *I*(*t*) as a function of the exposures for susceptible and infected for large enough populations (Fig. 7).

**Figure 7:**
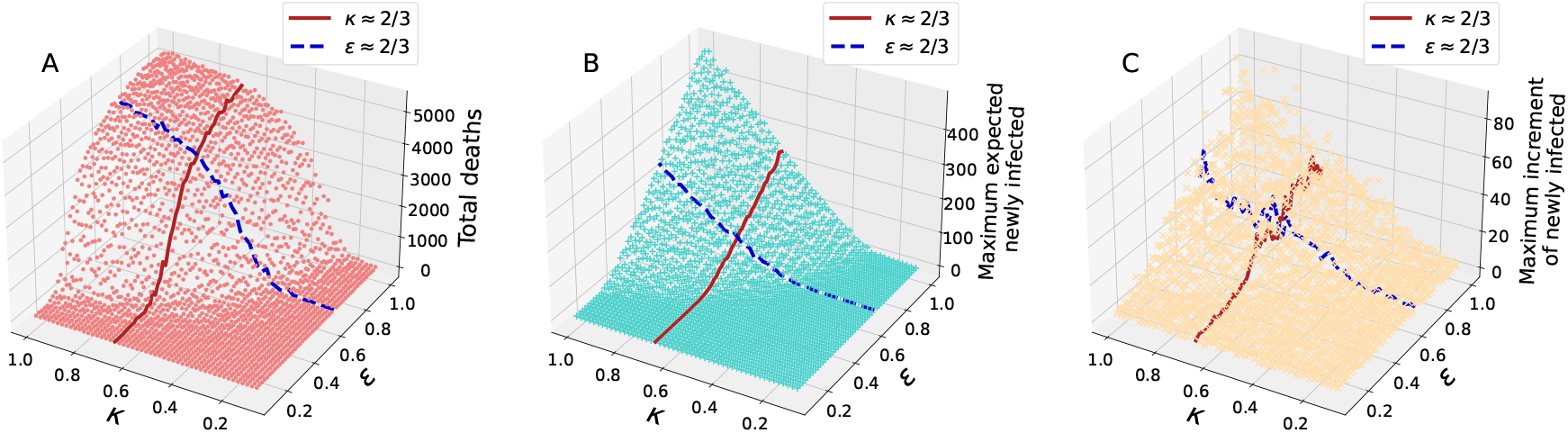
Quantities of interest that directly depend on *I*(*t*), as a result of varying the exposure for susceptible and infected (*ε* and *κ*) in the basic sED model. The solid line in red corresponds to the case *κ* = 2*/*3, and the dashed line in blue corresponds to the case *ε* = 2*/*3. (A) Total deaths, (B) maximum expected value of the newly infected individuals, and (C) maximum increment of the newly infected individuals between *t* and *t* + *h*.

In principle, it is possible to estimate the value of *p*^*h*^(*t, I*)*ε*(*t*), if data of new infections is available. For infectious diseases like COVID-19, or Ebola, if time is in minutes, or larger, and *h* is only a few seconds (*h <<* 1), then *p*_*I*_(*h*) ≈ 1, and the probability of infection given a contact with an infected individual is approximately *β κ*(*t*)*I*(*t*)*/T*(*t*), as it has been proposed for models like the classical SIR.

### 3.3 Case fatality ratios and the necessity of considering infections with different severity stages

The case-fatality ratios in the models just presented are very high with respect to what has been recorded for most epidemics, in spite of the fact that the times for recovery and death that were used are as recorded from data during the COVID-19 pandemic. At the heart of the matter is the fact that, during the first months of the COVID-19 pandemic, there were several studies reporting shorter hospitalisation times with fatal outcomes in comparison to recovery times (Alimohamadi et al., 2021; Porcheddu et al., 2020). This is the most important aspect in which the basic sED model (and the SIR as a particular case), falls short. How can we model several infectious stages that include disease, but simultaneously, have cases in which the time to dead, which involves passing through all possible stages (in common sense terms, such that infection cannot get worse unless it has become bad), to be shorter than the time it takes for an infected and sick individual to recover?

To address this issue, we propose an adaptation of the basic model that takes into account how the routing dynamics affect the sizes of the recovered and deceased subpopulations. The main idea is to take a more realistic description of the disease history of individuals into account. The extended model enables the possibility of reproducing both the macroscopic infection dynamics and the more detailed evolution of the recovered and deceased subpopulations (Fig. 8 and 9).

**Figure 8:**
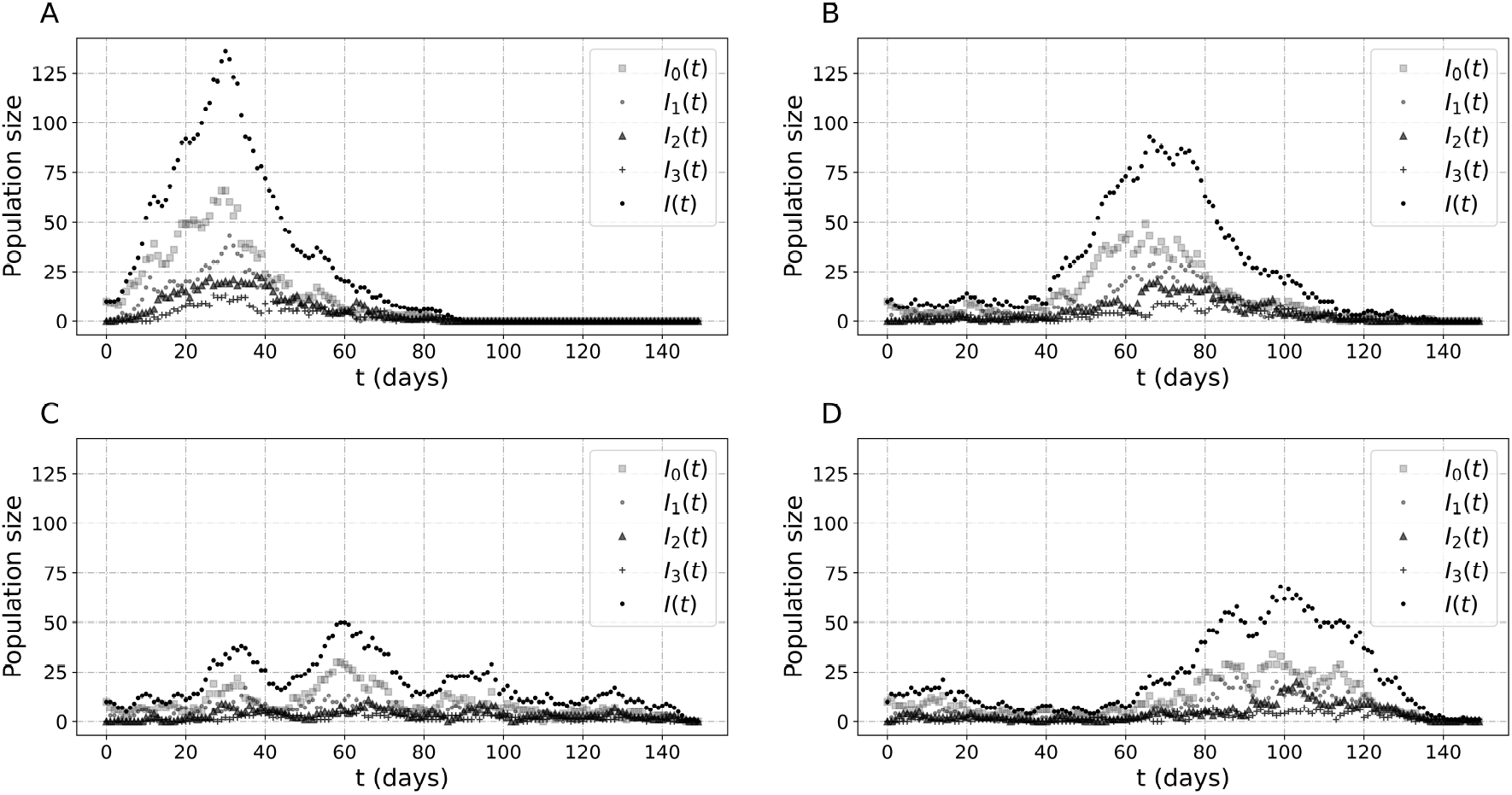
Different dynamics observed in the multistage sED. Realisations in all cases were simulated assuming the same total expected infection time with 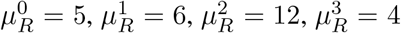 and 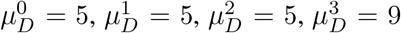 for the multistage sED. As before, the initial population size for these simulations is 1000.

**Figure 9:**
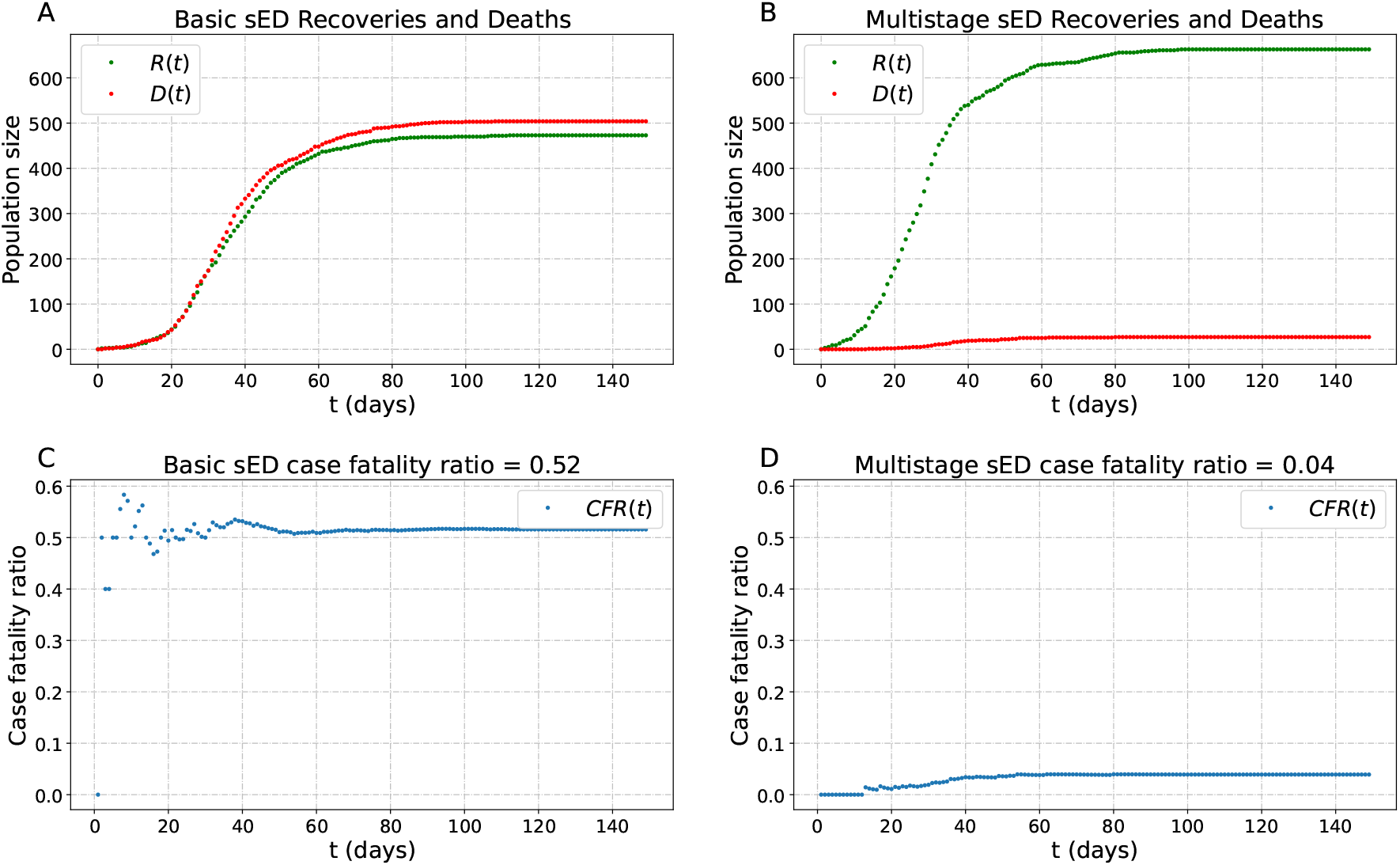
Case fatality ratios can be model more realistically by assuming multiple infection-related epidemiological stages. Epidemiological dynamics of multiple infection-related stages (A, C, respectively) *versus* a single infectious stage (B, D, respectively). Realisations in both cases were simulated assuming the same total expected infection time with *μ*_*R*_ = 27 and *μ*_*D*_ = 24 for the basic sED and 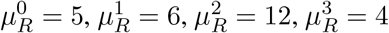, and 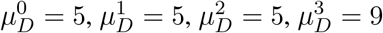 the multistage sED. As before, the initial population size for these simulations is 1000.

The main aspects addressed with the extended model include the following: The probability that an individual recovers eventually (equation (8)) depends on two quantities of statistical nature, *μ*_*R*_ and *μ*_*D*_, respectively representing the mean time spent by an individual in the infectious state and the mean time spent infected before death. However, equation (8) does not capture the complexity of the relation between epidemiological waiting times like *μ*_*R*_ and *μ*_*D*_, and the probability of having a good or a bad outcome. For example, the incubation time (the time between the initial inoculation and the emergence of symptoms) for some individuals may be very small, as they quickly develop an increasingly severe disease, significantly increasing the probability of death in comparison to other cases in which the disease is not as severe (Salinas-Escudero et al., 2020). Also, for some diseases the probability of recovery may decrease as individuals spend more days in a hospital (Faes et al., 2020; Salinas-Escudero et al., 2020). In addition, the participation of infected individuals in the chain of transmission may be reduced as the infection, and possibly the disease that it causes, progresses.

Naturally, assuming *μ*_*R*_ *> μ*_*D*_ for the basic sED model (and the SIR model) yields case-fatality ratios *D/*(*D* + *R*) > 1*/*2, which does not necessarily happen for most infectious diseases. In particular, for COVID-19 *μ*_*R*_ *< μ*_*D*_ and the CFR has been considerably smaller than 0.1 for all variants and all age groups (Bendavid et al., 2021; Cao et al., 2020; Luo et al., 2021). Thus, the most simple form of the sED model cannot yield small case-fatality ratios and simultaneously capture the macroscopic trends in the time course of *I*(*t*). By extension, the same happens with the SIR model.

#### 3.3.1 Infections with multiple states

Before developing the extension model, it is important to remark that the time period in which an individual is capable of transmitting the infection does not necessarily coincide with the manifestation of clinical symptoms. For instance, shedding of viable inoculum occurs during the incubation period for many infectious diseases. Alternatively, it is also possible that an infected person stops transmitting viable virions or other infection-causing pathogens before recovering (Li, 2010). These events depend strongly on the disease (Baron, 1964). Of interest, it could also be the case that individuals in advanced stages of infection do not participate in the chain of transmission because of being spatially isolated; *e*.*g*. due to hospitalisation, self isolation, or because of losing the ability to move.

To get a better epidemiological description of the initial infection and possibly the subsequent clinical stages, we model the increase in the severity of infection assuming that the time course of infection is divided in different stages that progress toward more severe illness, until possibly reaching the death state in the absence of recovery. In other words, at each infectious stage the individual either recovers, or advances to a more severe state. At the last infectious stage, the only possibilities are either recovery or death.

Assume that the sizes of the infected or ill populations are represented by *k* state variables {*I*_0_, …, *I*_*k−*1_}, with indices increasing according to the severity of the disease. The states do not necessarily represent clinical states. The size of the infected population is therefore *I* = *I*_0_ + … + *I*_*k−*1_. It is assumed here that a person may recover from any infection stage, and cannot become part of the subpopulation in an infection state without having been through all the infection states of less severity. By extension, an infected individual cannot die without having been through all the infection states. For instance, for COVID-19 it is reasonable to assume that there are at least four possible infectious states labelled as 0, 1, 2, and 3, with population sizes *I*_0_, *I*_1_, *I*_2_, *I*_3_, respectively representing an incubation period, followed by symptomatic stages *I*_1_, *I*_2_, and *I*_3_ of mild, middle, and severe stages, after which they could die (Fig. 8).

Let *hβ*_*i*_*κ*_*i*_(*t*) be the probability of having an effective transmission within a time window of size *h*, given a contact between a susceptible person and an individual in the stage *I*_*i*_. Then, by using the law of total probability counting over the partition on the different possible infectious subpopulations, the probability that a susceptible person becomes infected in a time window of size *h* is given by

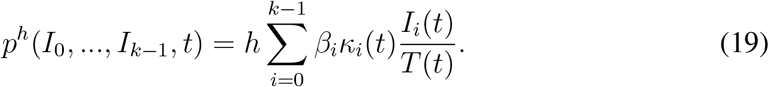

which generalises (18).

For the subsequent stages, in analogy with the Subsection 2.3.2, we can derive Geometrically distributed times for advancing or recovering to the next stage for each individual. The probabilities that a person in the *i*th infectious stage advances to the next stage or recovers, within a small time interval of length *h* are estimated by

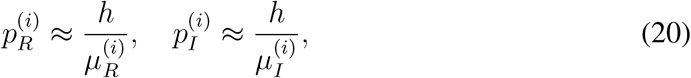

for *i* ∈ {0, …, *k* − 1}, where 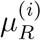 is the mean time that a patient spends in the *i*th infectious stage before recovering and 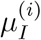 is the mean elapsed time that a patient spends in the *i*th infectious stage before moving forward to the next infectious stage. Accordingly, an individual remains infected in the *i*th stage with probability 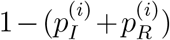. For computational and notational purposes, we can think the death stage as a stage labeled *k* with size *I*_*k*_, and use this extra index in equation (20). Therefore, a Multinomial approach can be derived in each stage, to compute the number of individual that advances or recover in each state to give a complete description of the dynamics for this extension. A system that extends equations (3)-(5) taking into account the different infectious stages can thus be written as

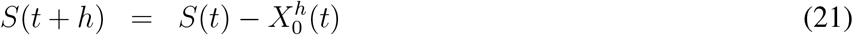

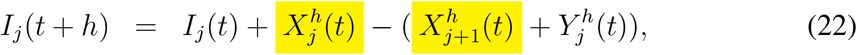

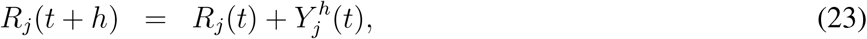

for *j* ∈ {0, …, *k* − 1}, with total counts for recovered and dead given by

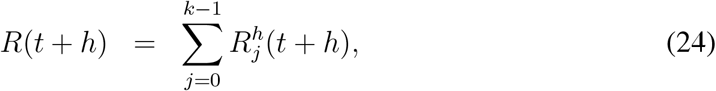

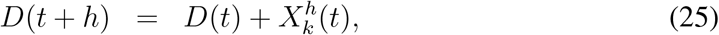

where *S*(*t*), *I*(*t*) = *I*_0_(*t*) + … + *I*_*k−*1_(*t*), *R*(*t*), and *D*(*t*) respectively represent the numbers of susceptible, infected, recovered, and dead individuals at time *t*. The sampling underlying the evolution is given by

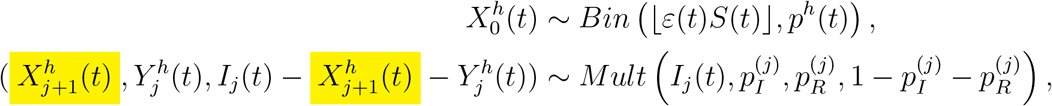

for *j* ∈ {0, 1, …, *k*}, assuming that all samples are independent. The *X*^*′*^*s* represent the quantity of people advancing through infectious stages, the last one being death, and the *Y* ^*′*^*s* represent the number of people recovering from the disease, at some given time.

The dynamics from this extension of the basic sED can be such that the total expected time before recovery is larger than the total expected time before death due to disease (Fig. 9), but such that the case fatality ratio is much smaller than 1/2 depending on the parameters of the model.

#### 3.3.2 Probability distribution of the CFR and the basic reproductive number

It is desirable to know the distribution of the total infected and recovered populations in order to characterise the distribution of the CFR in the multistage sED model. An approach to this problem is to use the continuous time Markov approximation obtained in Section 2.4 and to solve the asociated forward Kolmogorov equations. Those equations have been previously studied, see its relation with PDEs in Chalub and Souza (2011) and a numerical approximation in (Keeling and Ross, 2008), which corresponds to the continuous approximation for the basic sED model. In this section we derive an analogue of the basic reproduction number for the multistage sED model. Then, by means of simulations, we establish a probabilistic phase transition for the distribution of the (final) CFR determined by that number.

Recall that the basic reproduction number in our model is defined as the average number of secondary cases arising from an average primary case in an entirely susceptible population (Keeling and Rohani, 2008). In the basic SIR model the basic reproduction number determines whether there is an epidemic or not and arises from analysing when the infinitesimal increment of infected people is positive, *∂*_*t*_*y >* 0 (Allen et al., 2008).

Following last intuition, for the multistage sED model, we compute 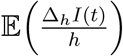, where Δ_*h*_*I*(*t*) = *I*(*t* + *h*) − *I*(*t*). First, let us observe that Δ_*h*_*I*(*t*) is given by

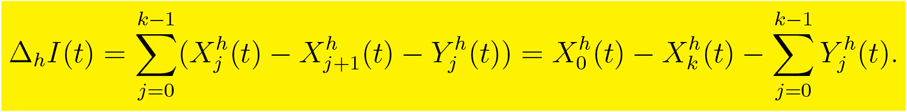

Therefore, we have that

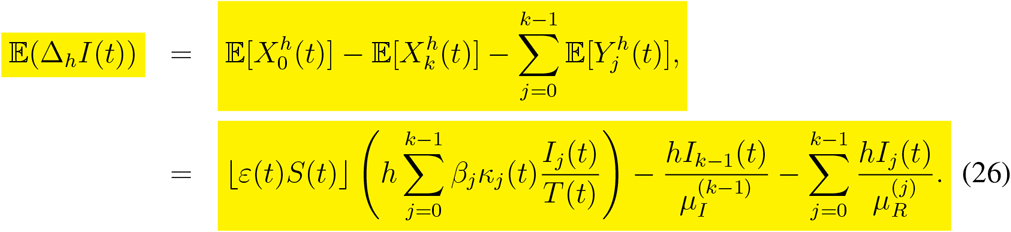

Note that at *t* = 0, assuming that the population is totally susceptible, there are not individuals in advanced stages of infection, that is *I*_*j*_(0) = 0 for every *j* ≠ 0. Hence,

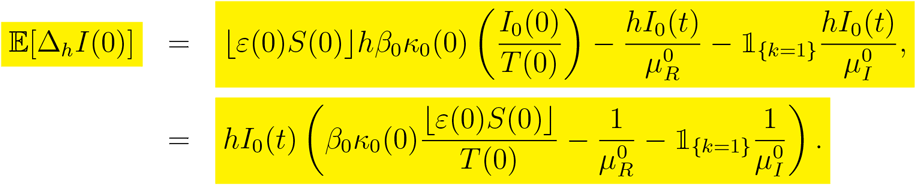

(The indicator function appears in last equation since, in the case of a single stage of infection, the flow of infected individuals who die should be taken into account for the total change in infected people). Therefore, we conclude that

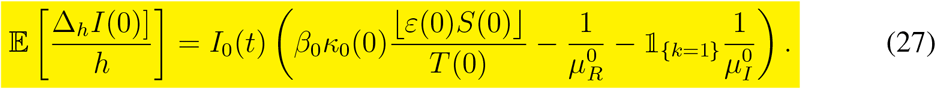

Consequently, in order for the expected rate of change in *I*(*t*) to be positive we obtain the condition

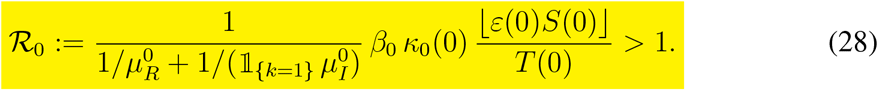

As a consequence, the average increment of the infected population with an almost entirely susceptible population will be larger than zero for ℛ_0_ > 1.

In the case of a unique stage of infection *k* = 1, if we consider total exposure of susceptible and infected individuals, *κ*(*t*) = *ε*(*t*) = 1, since 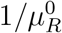 is the mean rate by time unit of infected people that recover and 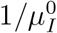 is the mean rate by time unit of infected people that die, we recover the usual R_0_ for a SIRD (Shringi et al., 2021).

There is a phase shift from a multimodal distribution to a unimodal distribution whose variance decreases (Fig. 10). This change occurs as *β*, the probability of infection given an infectious contact, increases. As a consequence, this change in the phase is tightly related with the basic reproductive number, ℛ_0_. Note there could be epidemic outbreaks even when the basic reproductive number for the multistage sED (equation (28)) is less than 1 (Fig. 10, panels A-D). In this case, the predominant mode accounts for all the realisations that ended without an epidemic outbreak and the lower mode accounts for all the realisations of epidemics, still with ℛ_0_ < 1. Additionally, given the change in phase between panel D and E of figure 10, when ℛ_0_ changes from being less than 1 to greater than 1.

**Figure 10:**
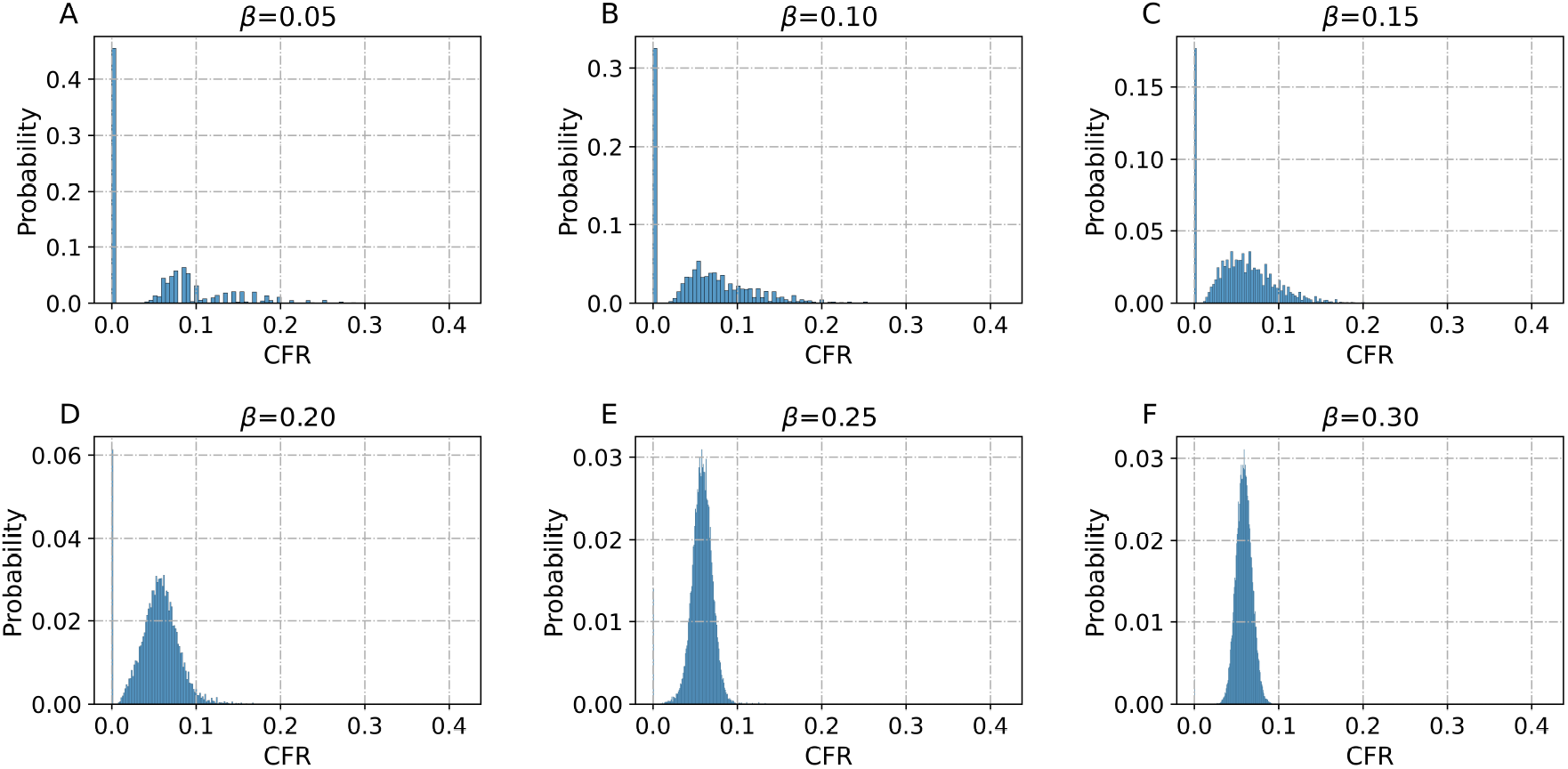
Final probability distribution for the case fatality ratio for different values of *β*. In each case 50,000 realisations were simulated with 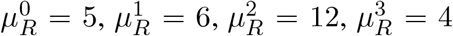, and 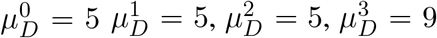 for the multistage sED. As before, the initial population size for these simulations is 1000. A phase shift from a multimodal distribution to a unimodal distribution can be observer when 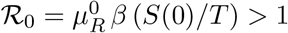, happening for *β >* 0.2.

The realisations discussed above show a qualitative change: the probability of observing outbreaks or whole epidemics is larger than 0 for ℛ_0_ < 1 and epidemics occur almost surely when ℛ_0_ > 1.

#### 3.3.3 Probability distribution of the final stage reached by an individual

In this section we illustrate some mathematical properties of the multistage extension of the sED model just presented. We focus on the maximal stage of infection or disease that a given individual may reach before recovering. The reason is that the last stage and the final clinical outcome is of interest for clinical and epidemiological purposes (Roger, 2011).

Consider the sED model with multiple infection stages *S, I*_0_, …, *I*_*k−*1_, *R, D* (equations (21)-(25)). Assume that one individual has been initially infected at some time *t*. By construction, the time spent infected and the maximal stage reached before recovering or dying will not depend on the (random) individual evolution.

First, using the same rationale as in the basic sED model, if an individual is in the *i*th stage of infection, the probability that the individual goes to the recovery state without passing to the next stage (without getting worse) is given by the routing probability of recovery 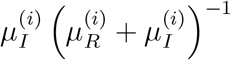 (see Subsection 2.3.2).

By considering the equations for each stage, assuming independence between the sampling processes at different stages, and considering only the expected times for each stage of infection, the probability that the infected individual reaches the stage *I*_*l*_ (*l* ≤ *k*) as maximal stage of infection or disease before recovering is given by

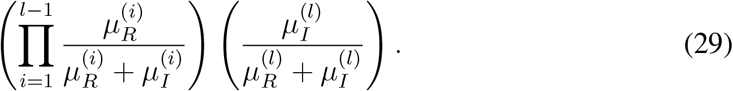

Using the same reasoning, the probability that a person dies is

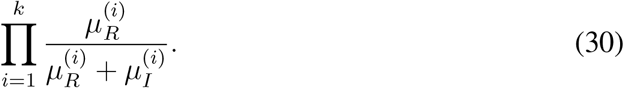

**Table 2:** Notation and values used in the multistage extension of the sED model (Li et al., 2020; Zhou et al., 2020).

| Symbol | Value | Units | Description |
| --- | --- | --- | --- |
| ine $\mu_R^0$ | [4.1,7.0] | days | Expected recovery time for asymptomatic people. |
| $\mu_R^1$ | [4.3,7.5] | days | Expected recovery time after first symptomatic stage. |
| $\mu_R^2$ | [9.0,15.0] | days | Expected recovery time spent after the second (mild) symptomatic stage. |
| $\mu_R^3$ | [2.0,9.0] | days | Expected recovery time spent in the third (severe) symptomatic stage. |
| $\mu_I^0$ | [4.1,7.0] | days | Expected incubation time. |
| $\mu_I^1$ | [4.3,7.5] | days | Expected time spent between the first and second symptomatic stages. |
| $\mu_I^2$ | [5.0,11.0] | days | Expected time spent in the second and third symptomatic stages. |
| $\mu_D = \mu_I^3$ | [4.0,12.0] | days | Expected time spent in the third (severe) symptomatic stage before death. |
| ine |  |  |  |

## 4 Discussion

We have presented a derivation of a model for the epidemiological dynamics of infectious diseases based on simple assumption: at any given time, individuals in any epidemiological stage are the subjects of a random sampling process in which they either remain in their epidemiological stage, or move to a different stage. Then we have distinguished two different types of sampling. In the first sampling type, individuals are chosen depending on an interaction with another subpopulation, as is the case during the infection process. In the second sampling type, sampling depends on waiting times. As a result, we have obtained a simple, easy to implement model derived from common sense rules, that takes into account different sources of randomness in the evolution of infectious diseases. The model works for small and large populations as well, and also, allows extensions to study the effects of specific factors that may become important determinants of the epidemics (e.g. exposure of infected *vs*. exposure of susceptible, single *vs*. multiple disease stages). While doing so, we explain how to derive a continuous time, continuously valued SIR model from the sED model, and in what circumstances do the classical SIR models work, and expose some of the problems that arise in extensions of the SIR models intended take into account deaths due to disease. We also show a few mathematical properties related to the expected times spent by an individual in different epidemiological stages. Of particular interest, we show how case fatality ratios in models with only one infectious stage would be larger than or equal to 1/2 unless the expected time for recovery is shorter than the expected time for death due to disease. We also show that realistic values for the CFR can be only be obtained in models with more than one infectious (or clinical) stage that allows percolation of the population through different infectious stages and recovery before reaching the last stage and dying.

### 4.1 Why do we need another model?

One issue of interest is the theoretical and practical impossibilities of using the continuous SIR model for small populations (equations (15)-(16)). However, there are many epidemics in which the populations of interest are small (e.g. small villages in Liberia during Ebola outbreaks). For large enough populations the dynamics of the deterministic, autonomous, and continuous SIR model can be compared to simulations of the dynamics obtained with the sED model after dividing the sED state variables by the population size (Fig. 5). In doing so, we show that simply adding another variable to the regular SIR deterministic dynamics to account for deceases leads to a contradiction, thereby showing that such a model would be conceptually ill-posed. Further, we illustrate the similarity between the dynamics of the sED model and the continuous state, deterministic model obtained in the limit for large populations. To do so, we show that the distance between the normalized 3-dimensional vector (*S*(*t*), *I*(*t*), *R*(*t*))*/T*(*t*) and its deterministic counterpart (*x*(*t*), *y*(*t*), *z*(*t*)) decreases robustly as the population size *T* increases (Fig. 5). In addition, our modelling paradigm also shows counter-intuitive situations. For instance, with our model it is possible to observe infectious outbreaks without epidemics, even for ℛ_0_ < 1. Similarly, we can observe outbreaks, and not whole epidemics, for ℛ_0_ > 1. Both situations allow the researcher a wider perspective about epidemics in comparison with deterministic models.

There are many possible extensions that could be implemented from the sED model. For instance, the assumption of having a homogeneous population can be easily dropped by allowing different probabilities of infection for more than one subpopulation. One easy way to do this is to use object oriented programming to create a generic class with sED dynamics for a single homogeneous population. Then it is possible to simulate the joint dynamics of more than one homogeneous population by setting a meta-class that allows the interactions of several instances of the class for a single population (Supplementary Material 1). The need for better understanding of the epidemiology and immunology underlying different co-morbidities for COVID-19 and other infectious diseases has been repeatedly suggested by surveillance data, genetic profiling data (Velavan et al., 2021), CT-value readings in RT-PCR tests (Waudby-West et al., 2021), and serological testing (Toulis, 2020). The modelling scheme presented here could also be extended to take into account populations with different susceptibilities. For instance, it became clear very early during the COVID-19 pandemic that overweight or obese people were at a larger risk of having a severe or lethal outcomes upon infection (Busetto et al., 2020; Hamer et al., 2020; Popkin et al., 2020). Another possible extension could aim to model epidemics with different mechanisms of infection.

The modelling scheme presented here allows the construction of extensions of the basic sED model to address these and other issues using data, which opens the possibility of adding a layer of knowledge and understanding that would potentially have a positive impact on decision making policies, and help health care providers.

### 4.2 The effects of exposure of susceptible and infected on the epidemic dynamics

Infection processes occurring between pairs of individuals depend at least in three aspects: the potential effectiveness of the infected individuals in transmitting the infection, the vulnerability to infection of the susceptible individuals, and the physical exposure between them, typically occurring through physical contact. These aspects are very difficult to measure precisely, but they can be thought of as independent, and different indirect statistical measures can be used to produce estimations for them. Examples of these include viral load as an indicator of infectivity potential (Jones et al., 2021; Marc et al., 2021); existence of comorbidities to measure vulnerability (MK et al., 2021); presence of antibodies in an individual as indicators of infection severity and history of infections (Legros et al., 2021); mobility in cities as a proxy of physical exposure (Lu and Gan, 2022), especially in consideration of lockdowns and social distance and public health measures (Wu et al., 2021). The sED model allows us to explicitly separate exposition of the susceptible population and the exposure of people with the potential to infect others. This allows us to observe explicitly the different dynamics resulting from having drastic reductions in each factor, opening the possibility of establishing or discarding the efficacy of public health measures directed to the reduction of such factors. To model one specific scenario that can be thought of in the context of the COVID-19 pandemic, we chose values for the exposures of infected people, *κ* ≈ 2*/*3, to produce a conservative simulation of a scenario in which the proportion of asymptomatic infected people was similar to that observed for COVID-19 (red curves, Fig. 7, He et al. (2021)). From those simulations, it can be appreciated that the strategy to mitigate the epidemics by reducing the exposure of people in general could fail if the values of *ε* are not small enough. That is, the strategy does not work unless the reduction in exposure for susceptible individuals decreases dramatically. For instance, for *κ* = 2*/*3, at most 1/3 of the susceptible individuals or less should be exposed in order to have a drastic reduction of in prevalence, incidence, and the resulting deaths. The strategy in this case was estimated so that death cases did not enter the parts of the curve with larger slopes (Fig. 7). Note that the strategy for health care providers and health system decision makers could be focused on the rate of change of the incidence instead using the same analysis.

Sadly, our model also confirms empirical observations that can be obtained from data of cases produced by the different SARS-CoV-2 variants around the world, showing that a necessary condition for the reduction of cases is to limit exposure of the infected, almost independently of the exposure of those susceptible (compare the cases for any fixed *κ* and different values of *ϵ*, and then viceversa in figure 7 to see the contrast). In particular, this means that it is nearly impossible to to mitigate epidemics in which those that are infected, asymptomatic, and contagious are exposed at large. One possibility in these cases is to set up control measures via direct epidemiological tracing measures, such as surveillance in public transportation stations and hubs. Our model complements previously well known results and demonstrates in a very simple way why consideration of transmission mitigation strategies based on lock-downs and limited mobility (Arino and Van den Driessche, 2003, 2006) should not be taken lightly, at least in the absence of effective surveillance (Bendavid et al., 2021; Sood et al., 2020) aimed at reducing the exposure of infected people to the general population. The results we show also make a case for the consideration of other mitigation strategies based on non-massive isolation and focused protection (Bhattacharya et al., 2021; Kullforff et al., 2021) that would also avoid economical and sociological harm imposed by lock-downs (Foster, 2020).

### 4.3 Infection vs infectivity

The sED model can describe some aspects concerning the infectivity and lethality of a disease that cannot be directly explained by the force of infection and the basic reproduction number. For a fact, but also for epidemiological purposes, the infection time may not necessarily be equal to the time interval in which a person is infectious. For instance, an infected person during the incubation period, or already in the process of clearing the virus, may have a sufficiently low viral load such that the person is effectively not infectious (Folgueira et al., 2021). Similarly, the infection time may include a period of symptoms that may be severe enough to reduce the exposure of the infected person (e.g. hospitalisation or self isolation), potentially decreasing the availability of inoculum shedding for other people to get infected. The sED model can be easily adjusted to study the case in which infectivity periods last less than the (clinical) infection time (and viceversa). Further, it is also possible to include decreased levels of exposure by the infected; we study those cases in a companion paper dedicated to study COVID-19.

Of note, hypothesis in the sED model that infection depends on the exposure of those infected in environments where susceptibles may be inoculated applies also to infections that depend on non-human vectors like dengue. This is justified by recent data from dengue outbreaks and endemicity in densely populated urban areas (Falcón-Lezama et al., 2017; Villabona-Arenas et al., 2016). Consider the case in which there is one female mosquito per building in an apartment complex. The radius of interaction of the mosquito can easily be 25 meters, without taking into account vertical interactions. This means that one case of dengue in one building could result in many more cases, without the need of an abundant population of mosquitoes. The sED model uses simple parameters that can, nevertheless, capture such complex infection processes. Those parameters can be calibrated using the statistical information available. Then the model can be used to analyse qualitatively different scenarios depending on those parameters. In particular, the multistage sED model allows to divide the process of infection using different layers related to different possible mechanisms of infection and transmission of the diseased studied.

#### 4.3.1 Multiple stages for infection and illness progression

As pointed out earlier, an adequate scheme to model the evolution of many diseases, should be one in which classification of infection states is constructed according to the physiological changes of the individual, whether or not they depend on clinical assessment. The clinical stages associated with infections and disease due to such infections can then also be considered. Note, however, that only taking clinical considerations into account may result in very inaccurate assessments with possibly undesired consequences. For instance, infections with SARS-CoV-2 cause a large percentage of asymptomatic cases (non-clinically detected), that in turn, has resulted in large increases in the incidence of cases that in some cities have overwhelmed the local health systems (Zhao et al., 2020). This is at large our motivation for thinking about the pathophysiological states of an individual during an infection as the base for our modelling scheme.

Constructing a model by taking the physiology related to the infection as a basis to classify different subpopulations may result in an inability to estimate parameters in an statistically meaningful way. However, the sED model is simple enough and easy to expand so that anyone could use it to explore qualitatively different outcomes of an epidemic, and identify underlying mechanisms guided by fitting real data (e.g. prevalence and incidence curves). That was the case leading to the extension of the basic sED model into the multistage model (Section 3.3), with which we show how CFRs with very low values can be obtained by simply introducing multiple, subsequent infection stages with the condition that individuals who die from the infection must go through all disease stages before dying. By means of exploration, the multistage sED model thus provided some insight about why it would not be possible to obtain low CFRs with shorter recovery intervals in comparison to hospitalisation times, as documented during the first months of the COVID-19 pandemic. That analysis also shows that it would not be possible to model COVID-19 with classical SIR dynamics unless the model is modified by stratifying the disease stages before death.

### 4.4 Limitations, future perspectives, and concluding remarks

We have presented a modelling scheme that allows a number of extensions to include particular situations. However, one limitation of our the modelling approach is that it does not distinguish between individuals in the same stage of infection. In particular, it ignores important characteristics for the dynamics, such as the individual viral load. Another limitation is that the model is memory-less: it is constructed by sampling and ignores the time dependence of factors, such as the individual recovery time, that could influence the dynamics.

One important factor to consider for possible extensions of the model is the variation in the behaviour of an individual’s immune system in the presence of microbes, and after an infection (Glück et al., 2005). The overall physiological state of the individual (Bryant and Curtis, 2013), and other genetic or epigenetic factors (Zhang and Cao, 2019) are important during the initial contact and the subsequent phases of infection.

For instance, recent evidence from the COVID-19 pandemic shows that clinical outcomes can be defined by factors such as the body fat ratio, which has been correlated to the capacity of an individual to produce inflammatory responses (Brojakowska et al., 2021). As a consequence, the individual profile that possibly includes details related to his/her immune status is essential to define possible courses of disease evolution. The hierarchy of models presented here can be used to model disease spread in small and large populations that may include individuals with different levels of susceptibility (work in progress).

It is worth mentioning that the concept of immunity, by name, is relatively ill posed, as large enough inoculations will almost surely result in symptomatic or even severe disease for people that already had contact with the infectious pathogen. In other words, reinfection is always a possibility for large enough inoculations, and possibly not so large ones, as shown by the massive number of COVID-19 reinfections. The severity of disease for reinfections can, in principle, be assumed to be mild or nonexistent. However, the recent COVID-19 pandemic provided with several counter-examples to that. In this regard, we are particularly interested in developing extensions addressing the study of epidemiological dynamics with reinfection, possibly involving different levels of severity in the reinfection outcomes (Montoya et al., 2013).

In this article we presented a new mathematical model to describe epidemiological dynamics. Its purpose is to describe some crucial aspects observed in the evolution of pandemics often not taken into account in the existing modelling literature, while simultaneously being intuitive, based on common sense arguments, and robust enough to be easily adapted to different particular features of diseases.

## Data Availability

All data produced in the present study are available upon reasonable request to the authors

## References

Yousef Alimohamadi, Habteyes Hailu Tola, Abbas Abbasi-Ghahramanloo, Majid Janani, and Mojtaba Sepandi. Case fatality rate of covid-19: a systematic review and meta-analysis. Journal of preventive medicine and hygiene, 62(2):E311, 2021.

Linda JS Allen. An introduction to stochastic epidemic models. In Mathematical epidemiology, pages 81–130. Springer, 2008.

Linda JS Allen. An introduction to stochastic processes with applications to biology. CRC press, 2010.

Linda J.S. Allen. A primer on stochastic epidemic models: Formulation, numerical simulation, and analysis. Infectious Disease Modelling, 2(2):128–142, 2017.

Linda JS Allen, Fred Brauer, Pauline Van den Driessche, and Jianhong Wu. Mathematical epidemiology, volume 1945. Springer, 2008.

Kristina Allers and Thomas Schneider. Ccr5δ32 mutation and HIV infection: basis for curative HIV therapy. Current Opinion in Virology, 14:24–29, 2015.

Abdulla M. Alsharhan. Survey of Agent-Based Simulations for Modelling COVID-19 Pandemic. Advances in Science, Technology and Engineering Systems Journal, 6(2): 439–447, 2021.

Dorothy Anderson and Ray Watson. On the spread of a disease with gamma distributed latent and infectious periods. Biometrika, 67(1):191–198, 1980.

Julien Arino and P Van den Driessche. A multi-city epidemic model. Mathematical Population Studies, 10(3):175–193, 2003.

Julien Arino and P Van den Driessche. Disease spread in metapopulations. Nonlinear Dynamics and Evolution Equations, Fields Inst. Commun, 48:1–13, 2006.

Samuel Baron. Mechanism of recovery from viral infection. Advances in virus research, 10:39–64, 1964.

Adhir K Basu. Introduction to Stochastic Process. Alpha Science Int’l Ltd., 2003.

Eran Bendavid, Bianca Mulaney, Neeraj Sood, Soleil Shah, Rebecca Bromley-Dulfano, Cara Lai, Zoe Weissberg, Rodrigo Saavedra-Walker, Jim Tedrow, Andrew Bogan, et al. Covid-19 antibody seroprevalence in santa clara county, california. International journal of epidemiology, 50(2):410–419, 2021.

Jay Bhattacharya, Sunetra Gupta, and Martin Kulldorff. Focused protection: The middle ground between lockdowns and “let it rip”, 2021.

Keith R Bissett, Jose Cadena, Maleq Khan, and Chris J Kuhlman. Agent-based computational epidemiological modeling. Journal of the Indian Institute of Science, pages 1–25, 2021.

Julie Boucau, Caitlin Marino, James Regan, Rockib Uddin, Manish C. Choudhary, James P. Flynn, Geoffrey Chen, Ashley M. Stuckwisch, Josh Mathews, May Y. Liew, Arshdeep Singh, Taryn Lipiner, Autumn Kittilson, Meghan Melberg, Yijia Li, Rebecca F. Gilbert, Zahra Reynolds, Surabhi L. Iyer, Grace C. Chamberlin, Tammy D. Vyas, Marcia B. Goldberg, Jatin M. Vyas, Jonathan Z. Li, Jacob E. Lemieux, Mark J. Siedner, and Amy K. Barczak. Duration of Shedding of Culturable Virus in SARS-CoV-2 Omicron (BA.1) Infection. New England Journal of Medicine, 387(3):275–277, 2022.

Teun Bousema, Lucy Okell, Ingrid Felger, and Chris Drakeley. Asymptomatic malaria infections: detectability, transmissibility and public health relevance. Nature Reviews Microbiology, 12(12):833–840, 2014.

Fred Brauer, Carlos Castillo-Chavez, and Zhilan Feng. Mathematical models in epidemiology, volume 32. Springer, 2019.

Tom Britton. Stochastic epidemic models: A survey. Mathematical Biosciences, 225(1): 24–35, 2010.

Agnieszka Brojakowska, Abrisham Eskandari, Malik Bisserier, Jeffrey Bander, Venkata Naga Srikanth Garikipati, Lahouaria Hadri, David A Goukassian, and Kenneth M Fish. Comorbidities, sequelae, blood biomarkers and their associated clinical outcomes in the mount sinai health system covid-19 patients. PLoS One, 16(7):e0253660, 2021.

Penelope A Bryant and Nigel Curtis. Sleep and infection: no snooze, you lose? The Pediatric infectious disease journal, 32(10):1135–1137, 2013.

Luca Busetto, Silvia Bettini, Roberto Fabris, Roberto Serra, Chiara Dal Pra, Pietro Maffei, Marco Rossato, Paola Fioretto, and Roberto Vettor. Obesity and covid-19: an italian snapshot. Obesity, 28(9):1600–1605, 2020.

Y. Cao, A. Hiyoshi, and S. Montgomery. Covid-19 case-fatality rate and demographic and socioeconomic influencers: worldwide spatial regression analysis based on country-level data. BMJ Open, 3(10):(11):e043560, 2020.

Fabio ACC Chalub and Max O Souza. The sir epidemic model from a pde point of view. Mathematical and Computer Modelling, 53(7-8):1568–1574, 2011.

Angela S Clem. Fundamentals of Vaccine Immunology. Journal of global infectious diseases, 3(1):73, 2011.

Cheryl Cohen, Jackie Kleynhans, Jocelyn Moyes, Meredith L McMorrow, Florette K Treurnicht, Orienka Hellferscee, Azwifarwi Mathunjwa, Anne von Gottberg, Nicole Wolter, Neil A Martinson, et al. Asymptomatic transmission and high community burden of seasonal influenza in an urban and a rural community in South Africa, 2017–18 (PHIRST): a population cohort study. The Lancet Global Health, 9(6):e863–e874, 2021.

Richard Dicker C., Fátima Coronado, Denise Koo, and R. Gibson Parrish. Principles of epidemiology in public health practice; an introduction to applied epidemiology and biostatistics. Self-study course. U.S. Department of Health and Human Services, Centers for Disease Control and Prevention (CDC), Office of Workforce and Career Development, 3rd edition, 2006.

Philipp Doenges, Thomas Götz, Tyll Krueger, Karol Niedzielewski, Viola Priesemann, and Moritz Schaefer. SIR-Model for Households. Arxiv, 2301.04355, 2023.

Denise L Doolan, Carlota Dobaño, and J Kevin Baird. Acquired immunity to malaria. Clinical microbiology reviews, 22(1):13–36, 2009.

AM El-Sayed, P Scarborough, L Seemann, and S. Galea. Social network analysis and agent-based modeling in social epidemiology. Epidemiologic Perspectives & Innovations, 9(1), 2012.

C. Faes, S. Abrams, D. Van Beckhoven, G. Meyfroidt, E. Vlieghe, and N. Hens. Time between Symptom Onset, Hospitalisation and Recovery or Death: Statistical Analysis of Belgian COVID-19 Patients. International Journal of Environmental Research and Public Health, 17(20):7560, 2020.

Jorge Abelardo Falcón-Lezama, René Santos-Luna, Susana Román-Pérez, Ruth Aralí Martínez-Vega, Marco Arieli Herrera-Valdez, Ángel Fernando Kuri-Morales, Ben Adams, Pablo Antonio Kuri-Morales, Malaquías López-Cervantes, and José Ramos-Castañeda. Analysis of spatial mobility in subjects from a dengue endemic urban locality in Morelos State, Mexico. PloS one, 12(2):e0172313, 2017.

Z. Feng, D. Xu, and H. Zhao. Epidemiological Models with Non-Exponentially Distributed Disease Stages and Applications to Disease Control. Bull. Math. Biol., 69: 1511–1536, 2007.

Frank Fenner, Peter A Bachmann, E Paul J Gibbs, Frederick A Murphy, Michael J Studdert, and David O White. Viral replication. Veterinary Virology, page 55, 1987.

Maria Dolores Folgueira, Joanna Luczkowiak, Fatima Lasala, Alfredo Perez-Rivilla, and Rafael Delgado. Prolonged sars-cov-2 cell culture replication in respiratory samples from patients with severe covid-19. Clinical Microbiology and Infection, 27(6):886–891, 2021.

Gigi Foster. Early estimates of the impact of covid-19 disruptions on jobs, wages, and lifetime earnings of schoolchildren in australia. Australian Journal of Labour Economics, 23(2):129–152, 2020.

Giuseppe Gaeta. A simple sir model with a large set of asymptomatic infectives. arXiv preprint arXiv:2003.08720, 2020.

Monica Gandhi, Deborah S Yokoe, and Diane V Havlir. Asymptomatic transmission, the Achilles’ heel of current strategies to control Covid-19, 2020.

Luis F. García. Immune Response, Inflammation, and the Clinical Spectrum of COVID-19. Frontiers in Immunology, 11:1441, 2020.

Charles P Gerba. Environmentally transmitted pathogens. In Environmental microbiology, pages 445–484. Elsevier, 2009.

Thomas Glück, Bernhard Kiefmann, Mathias Grohmann, Werner Falk, Rainer H Straub, and Jürgen Schölmerich. Immune status and risk for infection in patients receiving chronic immunosuppressive therapy. The Journal of Rheumatology, 32(8):1473–1480, 2005.

Laura Grange, Etienne Simon-Loriere, Anavaj Sakuntabhai, Lionel Gresh, Richard Paul, and Eva Harris. Epidemiological risk factors associated with high global frequency of inapparent dengue virus infections. Frontiers in Immunology, 5:280, 2014.

Major Greenwood. On the statistical measure of infectiousness. Epidemiology & Infection, 31(3):336–351, 1931.

Priscilla E Greenwood and Luis F Gordillo. Stochastic epidemic modeling. In Mathematical and statistical estimation approaches in epidemiology, pages 31–52. Springer, 2009.

Mark Hamer, Catharine R Gale, Mika Kivimäki, and G David Batty. Overweight, obesity, and risk of hospitalization for covid-19: A community-based cohort study of adults in the united kingdom. Proceedings of the National Academy of Sciences, 117(35): 21011–21013, 2020.

W.S. Hart, L.F.R. Hochfilzer, N.J. Cunniffe, H. Lee, H. Nishiura, and R.N. Thompson. Accurate forecasts of the effectiveness of interventions against ebola may require models that account for variations in symptoms during infection. Epidemics, 29:100371, 2019.

WS Hart, PK Maini, CA Yates, and RN Thompson. A theoretical framework for transitioning from patient-level to population-scale epidemiological dynamics: influenza a as a case study. Journal of the Royal Society Interface, 17(166):20200230, 2020.

Jingjing He, Yifei Guo, Richeng Mao, and Jiming Zhang. Proportion of asymptomatic coronavirus disease 2019: A systematic review and meta-analysis. Journal of medical virology, 93(2):820–830, 2021.

Xi He, Eric HY Lau, Peng Wu, Xilong Deng, Jian Wang, Xinxin Hao, Yiu Chung Lau, Jessica Y Wong, Yujuan Guan, Xinghua Tan, et al. Temporal dynamics in viral shedding and transmissibility of COVID-19. Nature medicine, 26(5):672–675, 2020.

Carlos Ignacio Herrera-Nolasco. Modelo epidemiológico probabilista con extensión a grupos metapoblacionales. Master’s thesis, Facultad de Ciencias, UNAM, Av. Universidad 3000, Circuito Exterior SN, 7 2020.

T Déirdre Hollingsworth, Roy M Anderson, and Christophe Fraser. Hiv-1 transmission, by stage of infection. The Journal of infectious diseases, 198(5):687–693, 2008.

Elizabeth Hunter, Brian Mac Namee, and John D. Kelleher. A taxonomy for agent-based models in human infectious disease epidemiology. Journal of Artificial Societies and Social Simulation, 20(3):2, 2017.

Terry C. Jones, Guido Biele, Barbara Mühlemann, Talitha Veith, Julia Schneider, Jörn Beheim-Schwarzbach, Tobias Bleicker, Julia Tesch, Marie Luisa Schmidt, Leif Erik Sander, Florian Kurth, Peter Menzel, Rolf Schwarzer, Marta Zuchowski, Jörg Hofmann, Andi Krumbholz, Angela Stein, Anke Edelmann, Victor Max Corman, and Christian Drosten. Estimating infectiousness throughout SARS-CoV-2 infection course. Science, 373(6551):eabi5273, 2021.

Matt J. Keeling and Pejman Rohani. Modeling Infectious Diseases in Humans and Animals. Princeton University Press, 2008. ISBN 9780691116174. URL http://www.jstor.org/stable/j.ctvcm4gk0.

Matthew James Keeling and Joshua V Ross. On methods for studying stochastic disease dynamics. Journal of the Royal Society Interface, 5(19):171–181, 2008.

WO Kermack and AG McKendrick. Contributions to the mathematical theory of epidemics. Proc Roy Soc Lond, 115:700–721, 1927.

M Kullforff, Sunetra Gupta, and Jay Bhattacharya. Great barrington declaration.(2020). URL https://gbdeclaration.org, x2021.

V. Legros, S. Denolly, and M. et al. Vogrig. A longitudinal study of SARS-CoV-2-infected patients reveals a high correlation between neutralizing antibodies and COVID-19 severity. Cell Mol Immunol, 18:318–327, 2021.

Michael Y. Li. An Introduction to Mathematical Modeling of Infectious Diseases, volume 2. Mathematics of Planet Earth, Springer, 2010.

Qun Li, Xuhua Guan, Peng Wu, Xiaoye Wang, Lei Zhou, Yeqing Tong, Ruiqi Ren, Kathy SM Leung, Eric HY Lau, Jessica Y Wong, et al. Early transmission dynamics in wuhan, china, of novel coronavirus–infected pneumonia. New England journal of medicine, 2020.

Alun L. Lloyd. Realistic distributions of infectious periods in epidemic models: Changing patterns of persistence and dynamics. Theoretical Population Biology, 60(1):59–71, 2001.

Huan Lu and Hongcheng Gan. Evaluation and prevention and control measures of urban public transport exposure risk under the influence of COVID-19—Taking Wuhan as an example. PLOS ONE, 17(6):1–15, 06 2022.

G. Luo, X. Zhang, H. Zheng, and D. He. Infection fatality ratio and case fatality ratio of COVID-19. Int J Infect Dis, 113:43–46, 2021.

Rose H. Manjili, Melika Zarei, Mehran Habibi, and Masoud H. Manjili. Covid-19 as an acute inflammatory disease. The Journal of Immunology, 205(1):12–19, 2020.

Aurélien Marc, Marion Kerioui, François Blanquart, Julie Bertrand, Oriol Mitja, Marc Corbacho-Monné, Michael Marks, and Jeremie Guedj. Quantifying the relationship between SARS-CoV-2 viral load and infectiousness. eLife, 10:e69302, 2021.

Singh MK, Mobeen A, Chandra A, Joshi S, and Ramachandran S. A meta-analysis of comorbidities in COVID-19: Which diseases increase the susceptibility of SARS-CoV-2 infection? Comput Biol Med, 130:104219, 2021.

Magelda Montoya, Lionel Gresh, Juan Carlos Mercado, Katherine L Williams, Maria José Vargas, Gamaliel Gutierrez, Guillermina Kuan, Aubree Gordon, Angel Balmaseda, and Eva Harris. Symptomatic versus inapparent outcome in repeat dengue virus infections is influenced by the time interval between infections and study year. PLoS Negl Trop Dis, 7(8):e2357, 2013.

L. Perez and S. Dragicevic. An agent-based approach for modeling dynamics of contagious disease spread. International Journal of Health Geographics, 8(50):728–732, 2009.

Chayanon Phucharoen, Nichapat Sangkaew, and Kristina Stosic. The characteristics of covid-19 transmission from case to high-risk contact, a statistical analysis from contact tracing data. EClinicalMedicine, 27:100543, 2020.

H Vincent Poor. The maximum difference between the binomial and poisson distributions. Statistics & probability letters, 11(2):103–106, 1991.

Barry M Popkin, Shufa Du, William D Green, Melinda A Beck, Taghred Algaith, Christopher H Herbst, Reem F Alsukait, Mohammed Alluhidan, Nahar Alazemi, and Meera Shekar. Individuals with obesity and covid-19: A global perspective on the epidemiology and biological relationships. Obesity Reviews, 21(11):e13128, 2020.

Rossella Porcheddu, Caterina Serra, David Kelvin, Nikki Kelvin, and Salvatore Rubino. Similarity in case fatality rates (CFR) of COVID-19/SARS-CoV-2 in Italy and China. The Journal of Infection in Developing Countries, 14(02):125–128, 2020.

JA Quesada, A López-Pineda, VF Gil-Guillén, JM Arriero-Marín, F Gutiérrez, and C Carratala-Munuera. Incubation period of COVID-19: A systematic review and metaanalysis. Revista Clínica Española (English Edition), 2020.

VL Roger. Outcomes research and epidemiology: the synergy between public health and clinical practice. Circ Cardiovasc Qual Outcomes., 4(3):257–9, 2011.

G. Salinas-Escudero, M.F. Carrillo-Vega, V. Granados-García, and et al. A survival analysis of COVID-19 in the Mexican population. BMC Public Health, 20:1616, 2020.

B. Shayak and M. Sharma. A new approach to the dynamic modeling of an infectious disease. Math. Model. Nat. Phenom., 16:33, 2021. ISSN 1359-5938.

Sakshi Shringi, Harish Sharma, Pushpa Narayan Rathie, Jagdish Chand Bansal, and Atulya Nagar. Modified sird model for covid-19 spread prediction for northern and southern states of india. Chaos, Solitons & Fractals, 148:111039, 2021.

Eric Silverman, Umberto Gostoli, Stefano Picascia, Jonatan Almagor, Mark McCann, Richard Shaw, and Claudio Angione. Situating agent-based modelling in population health research. Emerging Themes in Epidemiology, 18(1):10, 2021.

Neeraj Sood, Paul Simon, Peggy Ebner, Daniel Eichner, Jeffrey Reynolds, Eran Bendavid, and Jay Bhattacharya. Seroprevalence of sars-cov-2–specific antibodies among adults in los angeles county, california, on april 10–11, 2020. Jama, 323(23):2425–2427, 2020.

P. Toulis. Estimation of Covid-19 prevalence from serology tests: A partial identification approach. J Econom., 220(1):193–213, 2020.

Henry C Tuckwell and Ruth J Williams. Some properties of a simple stochastic epidemic model of SIR type. Mathematical biosciences, 208(1):76–97, 2007.

J.J.A. van Kampen, D.A.M.C. van de Vijver, and P.L.A. et al. Fraaij. Covid-19: Does the infectious inoculum dose-response relationship contribute to understanding heterogeneity in disease severity and transmission dynamics? Nature Communications, 12:267, 2021.

Alexei Vazquez. Exact solution of infection dynamics with gamma distribution of generation intervals. Phys. Rev. E, 103:042306, Apr 2021.

Thirumalaisamy P Velavan, Srinivas Reddy Pallerla, Jule Rüter, Yolanda Augustin, Peter G Kremsner, Sanjeev Krishna, and Christian G Meyer. Host genetic factors determining COVID-19 susceptibility and severity. EBioMedicine, 72:103629, 2021.

Christian Julián Villabona-Arenas, Jessica Luana de Oliveira, Carla de Sousa-Capra, Karime Balarini, Celso Ricardo Theoto Pereira da Fonseca, and Paolo Marinho de Andrade Zanotto. Epidemiological dynamics of an urban Dengue 4 outbreak in São Paulo, Brazil. PeerJ, 4:e1892, 2016.

Rupert Waudby-West, Benjamin J. Parcell, Colin N.A. Palmer, Samira Bell, James D. Chalmers, and Moneeza K. Siddiqui. The association between SARS-CoV-2 RT-PCR cycle threshold and mortality in a community cohort. European Respiratory Journal, 58(1), 2021.

Shishi Wu, Rachel Neill, Chuan De Foo, Alvin Qijia Chua, Anne-Sophie Jung, Victoria Haldane, Salma M Abdalla, Wei-jie Guan, Sudhvir Singh, Anders Nordström, and Helena Legido-Quigley. Aggressive containment, suppression, and mitigation of COVID-19: lessons learnt from eight countries. BMJ, 375:e067508, 2021.

Qian Zhang and Xuetao Cao. Epigenetic regulation of the innate immune response to infection. Nature Reviews Immunology, 19(7):417–432, 2019.

Hongjun Zhao, Xiaoxiao Lu, Yibin Deng, Yujin Tang, and Jiachun Lu. Covid-19: asymptomatic carrier transmission is an underestimated problem. Epidemiology & Infection, 148, 2020.

Fei Zhou, Ting Yu, Ronghui Du, Guohui Fan, Ying Liu, Zhibo Liu, Jie Xiang, Yeming Wang, Bin Song, Xiaoying Gu, et al. Clinical course and risk factors for mortality of adult inpatients with COVID-19 in Wuhan, China: a retrospective cohort study. The Lancet, 395(10229):1054–1062, 2020.

